# Cohort Profile: PRECISE-DYAD: a prospective cohort study linking maternal and infant health trajectories in sub-Saharan Africa

**DOI:** 10.64898/2025.12.17.25342279

**Authors:** Marie-Laure Volvert, Milly Wilson, Robin Okello Owino, Angela Koech, Hawanatu Jah, Hannah Blencowe, Yahaya Idris, Onesmus Wanje, Isaak Mwaniki, Joseph Mutunga, Fatima Touray, Emily Mwadime, Anna Roca, Geoffrey Omuse, Rachel Craik, Fatoumata Kongira, Moses Mukhanya, Kalilu Bojang, Baboucar Njie, Marvine Ochieng, Umberto D’Alessandro, Grace Mwashigadi, Agnes M Mutua, Anne Rerimoi, Marleen Temmerman, Joseph Akuze, Melisa Martinez-Alvarez, Dorcas N Magai, Benjamin Barratt, Jing Li, Jaya Chandna, Melissa J. Gladstone, Amina Abubakar, Rachel M Tribe, Asma Khalil, Marianne Vidler, Tatenda Makanga, Tatiana Taylor Salisbury, Hiten D Mistry, Sophie E. Moore, Helen Nabwera, Veronique Filippi, Laura A Magee, Liberty Makacha, Lucilla Poston, Esperança Sevene, Peter von Dadelszen, the PRECISE-DYAD Network

## Abstract

**Purpose:** The PRECISE-DYAD study is a prospective observational cohort, designed to investigate health outcomes among mother-child pairs (dyads), over the first three years of life in two contexts from sub-Saharan Africa. The primary objective of the study was to explore the effects of selected placenta-related complications, such as pregnancy hypertension, fetal growth restriction, and preterm birth, on 1) child health and development, and 2) women’s health and well-being, including outcomes after stillbirth

**Participants:** The PRECISE-DYAD study enrolled women (and their children) originally recruited into the PRECISE regnancy cohort study in The Gambia and Kenya between July 2021 and April 2024. Participants were seen at 6 weeks to 6 months, 12 months, 24 months, and 36 months post-partum. Clinical and health data, including anthropometry and diet were collected for both mothers and children. Mother assessment included a cardiology assessment and collection of data about symptoms of COVID-19 infection. In a subset of participants, mothers were asked about their mental health, their health care costs during and after pregnancy, and experiences of care during labour and childbirth / delivery. Additonally, a personal environmental exposure assessement was performed for a subset of the cohort, by collecting air and water quality data alongside geographical, demographic, and behavioural factors. Child development was assessed using neurodevelopmental assessments, home environment evaluation, and quality of life measures. Biological samples were collected from mothers and children, processed promptly and biobanked locally. Sample data were entered into an OpenSpecimen database and linked to each individual, as well as to their corresponding social determinants and clinical data.

**Findings to date:** A total of 2,980 women and 2,909 children completed at least one PRECISE-DYAD study visit. The biorepository contains 108,897 biological samples from mothers and children. Baseline descriptive analysis of the cohort are reported here.

**Future plans:** Analysis of data and samples will include biomarker studies, social determinants of health, and epidemiological investigations. These analyses will explore how placenta-related complications and environmental exposures, such as nutrition and air quality, interact to shape maternal health, mental well-being, subsequent pregnancies, and mother-child interaction, as well as child growth and neurodevelopment through early childhood. Additional work will examine the biological pathways linking these exposures to outcomes and the impacts of caring for children with moderate-to-severe disabilities on maternal well-being. Findings will be disseminated through scientific publications, conference presentations, engagement with local stakeholders, and continued community outreach.

**Strengths and limitations:**

- This is a unique pregnancy-enrolled, population-based cohort with extensive social, clinical, and biological data, including biospecimens, collected across two geographically diverse settings in sub-Saharan Africa. Women were recruited at the time of booking for antenatal care, allowing early identification and longitudinal follow-up of those with placenta-related complications. The integration of PRECISE and PRECISE-DYAD data enables the comprehensive investigation of the drivers and impacts of placental disorders on maternal and child health, and outcomes related to the COVID-19 pandemic.
- Data were collected on women’s social and physical environments, including air quality, and water, sanitation, and hygiene (WASH) conditions. In-depth data were also gathered on children, with a focus on neurodevelopmental assessments. Consistent data collection procedures and standardised methodologies were used across both study sites.
- Extensive and sustained community engagement, including 108 sensitisation meetings with nearly 4,000 participants, enhanced trust, study understanding, and acceptability.
- A limitation of the study is the loss to follow-up of participants who relocated outside of the study area during pregnancy or after the child’s birth, or changed their contact details.
- A second limitation is that the Mozambique pregnancy cohort has provided only air quality data through PRECISE-DYAD, and has not been followed up otherwise at this time.

## Introduction

Despite global efforts to improve maternal and child health, low-and middle-income countries, including many in sub-Saharan Africa, bear a disproportionate burden of adverse pregnancy outcomes [1–3]. Pregnancy hypertension, fetal growth restriction (FGR), and stillbirth are linked to around 46,000 maternal deaths and 2.5 million fetal, neonatal, and infant deaths worldwide annually; more than half occur in sub-Saharan Africa [4]. While research from high-income countries has explored the long-term impact of maternal health and pregnancy complications on both maternal and child outcomes [5–7], the underlying mechanisms remain poorly understood. Maternal and child health in these regions is further affected by multiple co-exposures such as food insecurity and limited dietary diversity [6], endemic infectious diseases [7], poor air quality [8, 9] [8, 9], inadequate sanitation, and restricted access to healthcare [10–12]. Although studies have demonstrated that preterm birth, infections in pregnancy, and hypoxic-ischaemic encephalopathy are associated with poorer child health and neurodevelopmental delay, the mechanisms linking these pregnancy complications to long-term outcomes remain unclear. This is especially the case in sub-Saharan Africa and other low-resource settings, where the risk of developmental delay in childhood is elevated [13–15]. These overlapping exposures increase vulnerability, contributing to persistently high rates of maternal and perinatal morbidity and mortality, while the long-term consequences for mothers and children remain inadequately studied.

The PRECISE-DYAD study was established to address these gaps by building on the PRECISE project (PREgnancy Care Integrating Translational Science, Everywhere; https://precisenetwork.org/) [16–18]. PRECISE-DYAD is designed to investigate pathways of resilience and vulnerability that influence maternal and child outcomes following both pregnancy, including those complicated by hypertensive disorders, preterm birth, FGR, and stillbirth. The study has five key objectives: (1) to evaluate how placenta-related complications affect maternal health, including mental well-being, subsequent pregnancies, and mother–child interaction; (2) to examine how these complications influence child growth and neurodevelopment up to three years of age; (3) to explore the biological mechanisms underlying these outcomes; (4) to assess the impact of environmental exposures, such as nutrition and air quality; and (5) to determine how raising children with moderate-to-severe disabilities affects maternal health and well-being.

By integrating longitudinal clinical, epidemiological, and biological data with a comprehensive biorepository, PRECISE-DYAD provides a unique platform for advancing research on maternal and child health in two Sub-Saharan African countries, Kenya and The Gambia.

## Cohort description

### Study Setting

The PRECISE-DYAD study was undertaken in Kenya and The Gambia in collaboration with the Aga Khan University, Kenya (AKU) and the MRC Unit The Gambia (MRCG) at the London School of Hygiene and Tropical Medicine (LSHTM) [17].

In Kenya, the field research was conducted in two secondary level hospitals: Mariakani sub county Hospital (peri-urban) and Rabai sub county Hospital (rural). In The Gambia, the study was conducted in the Farafenni district, located close to The Gambia-Senegal border on the North Bank of the country. Field research took place at the Maternal Newborn Child and Adolescent Health clinic in Farafenni (an urban primary health centre [PHC]), the Farafenni General Hospital, and associated rural PHCs in Illiasa and Ngayen Sanjal.

### Patient and Public Involvement

Extensive community engagement has been integral to the PRECISE study since 2018, with research teams working in close collaboration with participating communities in Kenya and The Gambi a[17, 19]. During the PRECISE-DYAD study, this engagement continued with a strong emphasis on building trust and ensuring that the study was clearly understood by communities. Efforts were made to remain sensitive to community contexts, particularly in relation to the collection of biological samples. A total of 80 and 28 sensitisation meetings were held in Kenya and The Gambia, respectively, engaging more than 2,400 and 1,500 participants across both countries.

In addition,‘PRECISE-DYAD Open Days’ were organised once a month in the communities of participants. These events involved interactive learning activities and facilitated discussions on maternal and child health. Open days also offered an opportunity to gather feedback from participants about their experiences in the study and to identify any concerns or emerging issues. A total of 27 and 28 Open Days were held in Kenya and The Gambia, respectively, reaching 1,628 participants in kenya and 1,417 in the Gambia.

Towards the conclusion of the study, the PRECISE teams in Kenya and The Gambia conducted a series of dissemination meetings to discuss preliminary findings from both the main study and the air quality substudy. Once specific study results are published, participants will be informed through the PRECISE website (https://precisenetwork.org/) and will receive summaries in the form of infographics suitable for a non-specialist audience.

### Study Design

PRECISE-DYAD is an observational study, involving women and their children who were previously enrolled in the PRECISE study [16]. Women were recruited to the PRECISE cohort at their first antenatal care (ANC) visit between July 2019 and April 2022 and were invited to take part in the follow-up PRECISE-DYAD study between July 2021 and April 2024. This meant that participants entered DYAD at different stages; some joined early postpartum, others at later stages (see below)

The study protocol has been described in detail elsewhere [17, 18]. In brief, mothers and/or their infants were followed up to three years after birth. During this period, biological samples and clinical data were collected on both maternal and child health. In Kenya, a phone interview questionnaire was designed in order to collect key data between in-person visits; however, only 28 (<1%) participants were recruited through this method. A total of 2,980 women and 2,909 children were followed up and including 2,062 women and 2025 children in Kenya, and 918 women and 884 children in The Gambia.

### Study Population

Participants were enrolled at any point during the follow-up period when eligible for a PRECISE-DYAD visit (i.e., when they reached the appropriate age for that visit). A flow chart detailing the number of participants eligible, approached, and enrolled at each visit is presented in Figure 1, with country-specific information shown in Figures S1 (Kenya) and S2 (The Gambia). Participants enrolled in PRECISE (n=4122) and eligible to a specific study visit were called to participate to the PRECISE-DYAD study. Overall, a total of 2,864 participants (69.4%) were eligible for visit 1 (conducted 6 weeks to 6 months after birth). Of these, 2,205 women (76.9%) were successfully contacted, and 1,966 attended the visit (68.6% of those eligible). For visit 2, which took place at 12 months postpartum, 2,961 participants (71.8%) met the eligibility criteria. Among them, 2,233 (75.1%) were successfully approached, and 2,026 attended (68.4%). Eligibility and follow-up rates declined at later visits. At visit 3 (24 months postpartum), 3,137 women (76.1%) were eligible; 2,037 (64.9%) were contacted successfully, and 1,838 attended (58.5%). By visit 4 (36 months postpartum), 1,538 participants (37.3%) remained eligible. Of these, 905 (58.8%) were successfully approached, and 822 attended (53.4%). In some cases, only one member of the dyad was seen at a given visit. Mothers occasionally attended alone if their child was unwell and the visit could not be rescheduled, or in the event of child/pregnancy loss. Children attended with a caregiver if the biological mother was absent or in the event that she had died.

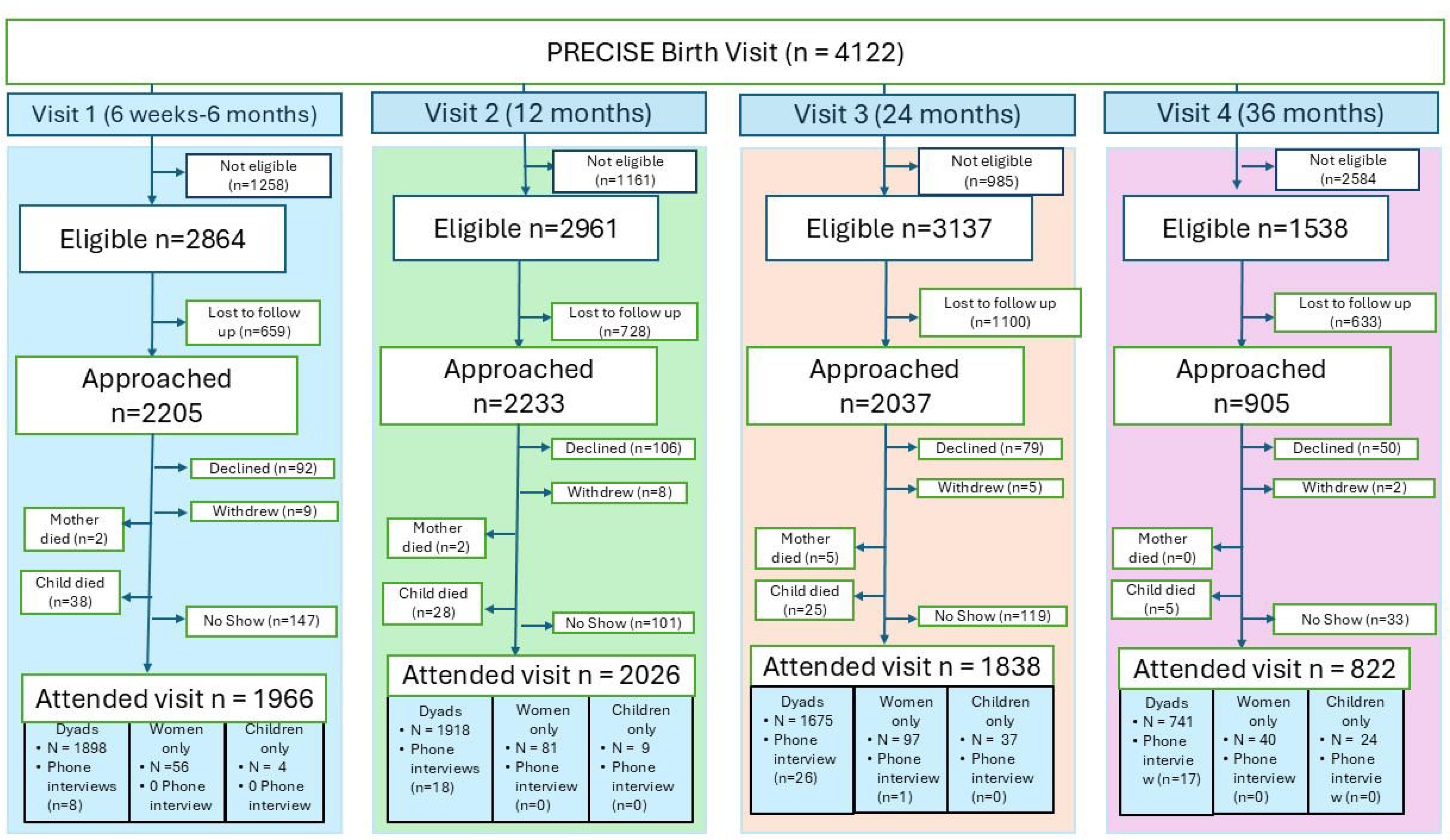

A flow diagram representing the follow up though the four study visits is described in Figure 2. Overall, 1,966 participants (65.9% of total participants who attended a visit) entered the study at visit 1, and 126 of these completed all four PRECISE-DYAD visits (6.4%). At visit 2, 428 participants (16.1%) joined the study, with 177 of them (41.3%) followed until visit 4. At visit 3, 436 participants (14.6%) were enrolled, and 316 of these (72.5%) continued through to visit 4. Finally, 163 participants (5.4%) were seen only at visit 4. Further details by countires are provided in Figures S3 (Kenya) and S4 (The Gambia).

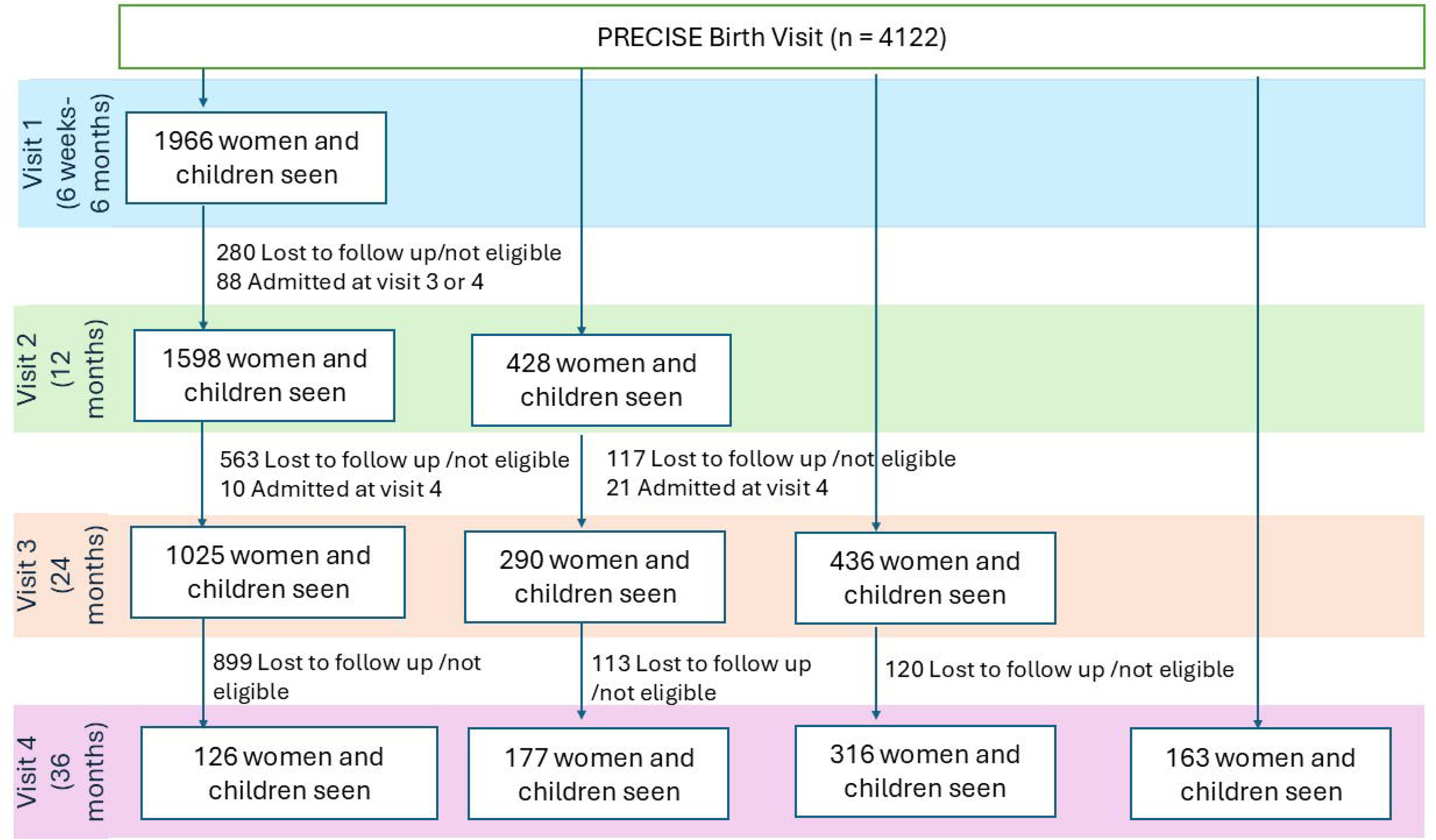

### Data Collection

Extensive clinical data were collected by trained research staff using Android tablets at participating healthcare facilities during each PRECISE-DYAD visit. Data collection was conducted through structured interviews, with biological samples collected in parallel. Details on the specific data and procedures followed at each visit are outlined in the study protocol described elsewhere [17, 18]. If key pregnancy-related information (e.g., date of birth, birthweight, sex of the baby, or maternal and neonatal outcomes), were missing, this information was obtained retrospectively during contact with participants for the PRECISE-DYAD study.

The air quality substudy assessed women personal air quality exposure using portable sensor packs (Dyson Technology Ltd, Malmesbury, UK), worn continuously over five days. These devices recorded levels of PM2.5, PM10, nitrogen dioxide, temperature, humidity, and mobility (via accelerometry and GPS). In addition to participants from Kenya and The Gambia, a subset of women in Mozambique were also included. The Mozambican women were recruited from two health facilities—Manhiça District Hospital and Xinavane Rural Hospital—as part of the PRECISE-HOME study.

### Data and Sample Management

Data management for the PRECISE-DYAD study was co-ordinated by a central team based at King’s College London, in collaboration with the recruiting hubs in Kenya and The Gambia. Each hub employed a Data Manager, responsible for maintaining the local database, including tasks such as data entry and cleaning. Local Data Managers ran queries to identify outliers, missing, or inconsistent data, while the central team provided monthly reports to the hubs, flagging any additional data issues. In addition, the co-ordinating team included a central Data Manager who oversaw the databases across both sites, handling tasks such as building and installing the database on local servers, running data queries, and extracting data for analysis. These procedures were implemented to ensure data accuracy.

All data and samples are owned by the country teams in which the participants resided during the study. Each country team maintains an updated version of their dataset, and any data-related queries are addressed directly through these teams. Updates are synchronised between the country-specific databases and the central database, which is currently hosted at King’s College London. Participants were assigned a unique study identification number used consistently across both the social determinants, clinical, and laboratory datasets. Identifiable personal information was retained exclusively within the country of data collection and was not shared externally.

Clinical data were collected using electronic data capture (EDC) on the ODK-X platform [20], via tablets during study visits. Database incorporated built-in validation rules and programming logic to implement skip logic and cross-validation, along with range limits for certain fields to reduce data entry errors.

An OpenSpecimen platform [21], was used for laboratory information management. The platform was configured with validation features to minimise errors during data collection and included an offline data collection tool for situations with limited internet connectivity. Data collected offline were uploaded once an internet connection was restored.

### Findings to Date

#### Participant demographics

The baseline characteristics of participating women who came to at least one DYAD visit are summarised in Table 1 and are shown with the demographic profiles of Kenyan and the Gambian and pregnancy cohorts also described in Craik et al., 2025 [16]. A total of 2952 participants were seen in person at least once during PRECISE-DYAD study and 28 particpants had a phone interview (only in Kenya). On average, women came for their first visit 5.1 months after delivery (IQR: 3.2, 12.4) with a minimum of 1.4 months after delivery and maximum of 37.9 months. This wide variability is due to the eligibility period for some participants after birth. A detailed description of participant characteristics at each visit is available in Table S1.

**Table 1.** Maternal demographic information of participants (from Kenya and The Gambia) originally recruited into the PRECISE study and subsequently followed up via the PRECISE-DYAD study. This table summarises demographic and clinical characteristics of women retained in the DYAD follow-up. Data from the original PRECISE pregnancy cohort are included for comparison, allowing assessment of differences between participants who continued in the postnatal follow-up and the broader cohort from which they were drawn. IQR Inter Quartile Range

|  | The Gambia |  | Kenya |  | Both countries |  |
| --- | --- | --- | --- | --- | --- | --- |
|  | PRECISE<br>Pregnancy | PRECISE-DYAD<br>(first visit attended) | PRECISE<br>Pregnancy | PRECISE-DYAD<br>(first visit attended) | PRECISE<br>Pregnancy | PRECISE-DYAD<br>(first visit attended) |
| **Number of participants (total)** | 1356 | 918 | 3670 | 2062 | 5026 | 2980 |
| Number of participants (in person) | - | 918 | - | 2034 | - | 2952 |
| Number of phone interview | - | NA | - | 28 | - | 28 |
| **Maternal age, median (IQR)** | 26.0<br>(22.0, 31.0) | 28.0<br>(24.0, 33.0) | 26.0<br>(23.0, 31.0) | 28.0<br>(24.0, 33.0) | 26.0<br>(22.0, 31.0) | 28.0<br>(24.0, 33.0) |
| **Age category, year, N(%)** |  |  |  |  |  |  |
| 15-19 | 169 (12.5) | 42 (4.6) | 258 (7.0) | 63 (3.1) | 427 (8.5) | 105 (3.5) |
| 20-24 | 359 (26.5) | 227 (24.7) | 1150 (31.3) | 522 (25.3) | 1509 (30.0) | 749 (25.1) |
| 25-29 | 386 (28.5) | 258 (28.1) | 1060 (28.9) | 644 (31.2) | 1446 (28.8) | 902 (30.3) |
| 30-34 | 245 (18.1) | 210 (22.9) | 747 (20.4) | 481 (23.3) | 992 (19.7) | 691 (23.2) |
| 35-39 | 134 (9.9) | 118 (12.9) | 747 (20.4) | 270 (13.1) | 496 (9.9) | 388 (13.0) |
| 40-44 | 47 (3.5) | 54 (5.9) | 81 (2.2) | 69 (3.3) | 128 (2.5) | 123 (4.1) |
| 45-49 | 4 (0.3) | 5 (0.5) | 5 (0.1) | 9 (0.4) | 9 (0.2) | 14 (0.5) |
| 50+ | 0 (0.0) | 1 (0.1) | 0 (0.0) | 1 (0.1) | 0 (0.0) | 2 (0.1) |
| missing | 12 (0.9) | 3 (0.3) | 7 (0.2) | 3 (0.1) | 19 (0.4) | 6 (0.2) |
| **Marital status, N(%)** |  |  |  |  |  |  |
| Never married (or single) | 24 (1.8) | 19 (2.1) | 223 (6.1) | 122 (5.9) | 247 (4.9) | 141 (4.7) |
| Married/ Co-habiting | 1318 (97.2) | 895 (97.5) | 3361 (91.6) | 1910 (92.6) | 4679 (93.1) | 2805 (94.1) |
| Separated/Divorced | 6 (0.4) | 4 (0.4) | 55 (1.5) | 24 (1.2) | 61 (1.2) | 28 (0.9) |
| Widowed | 0 (0.0) | 0 (0.0) | 8 (0.2) | 4 (0.2) | 8 (0.2) | 4 (0.1) |
| Missing | 8 (0.6) | 0 (0.0) | 23 (0.6) | 2 (0.1) | 31 (0.6) | 2 (0.1) |
| **Education, N(%)** |  |  |  |  |  |  |
| None | 851 (62.8) | 580 (63.3) | 341 (9.3) | 153 (7.4) | 1192 (23.7) | 733 (24.6) |
| Primary | 224 (16.5) | 152 (16.6) | 1932 (52.7) | 1076 (52.2) | 2156 (42.9) | 1228 (41.2) |
| Secondary | 214 (15.8) | 146 (15.9) | 945 (25.7) | 567 (27.5) | 1159 (23.1) | 713 (23.9) |
| Higher | 58 (4.3) | 40 (4.4) | 428 (11.7) | 264 (12.8) | 486 (9.7) | 304 (10.2) |
| Missing | 9 (0.7) | 0 (0.0) | 24 (0.7) | 2 (0.1) | 33 (0.7) | 2 (0.1) |
| **Occupation, N(%)** |  |  |  |  |  |  |
| Housewife | 1188 (87.6) | 808 (88.0) | 2007 (54.7) | 1091 (52.9) | 3195 (63.6) | 1899 (63.7) |
| Student | 13 (1.0) | 9 (1.0) | 76 (2.1) | 40 (1.9) | 89 (1.8) | 49 (1.6) |
| Professional | 21 (1.5) | 13 (1.4) | 261 (7.1) | 161 (7.8) | 282 (5.6) | 174 (5.8) |
| Factory | 0 (0.0) | 0 (0.0) | 88 (2.4) | 49 (2.4) | 88 (1.8) | 49 (1.6) |
| Large-scale agriculture | 11 (0.8) | 6 (0.7) | 4 (0.1) | 0 (0.0) | 15 (0.3) | 6 (0.2) |
| Market trader | 50 (3.7) | 34 (3.7) | 354 (9.7) | 226 (11.0) | 404 (8.0) | 260 (8.7) |
| Business | 0 (0.0) | 0 (0.0) | 348 (9.5) | 200 (9.7) | 348 (6.9) | 200 (6.7) |
| Informal - Employment | 0 (0.0) | 0 (0.0) | 471 (12.8) | 269 (13.0) | 471 (9.4) | 269 (9.0) |
| Other | 64 (4.7) | 48 (5.2) | 35 (1.0) | 22 (1.1) | 99 (2.0) | 70 (2.3) |
| Missing | 9 (0.7) | 0 (0.0) | 26 (0.7) | 4 (0.2) | 35 (0.7) | 4 (0.1) |
| **Returned to work/school since giving birth , N(%)** | - | 268 (29.2) | - | 756 (36.7) |  | 1024 (34.4) |
| **Religion, N(%)** |  |  |  |  |  |  |
| Muslim | 1340 (98.8) | 912 (99.3) | 1425 (38.8) | 782 (37.9) | 2765 (55.0) | 1694 (56.8) |
| Christian | 8 (0.6) | 6 (0.7) | 2209 (60.2) | 1271 (61.6) | 2217 (44.1) | 1277 (42.9) |
| Other | 0 (0.0) | 0 (0.0) | 12 (0.3) | 7 (0.3) | 12 (0.2) | 7 (0.2) |
| Missing | 8 (0.6) | 0 (0.0) | 24 (0.7) | 2 (0.1) | 32 (0.6) | 2 (0.1) |
| **Household composition** |  |  |  |  |  |  |
| Total Number of people in the household (IQR) | - | 13.0 (9.0, 18.0) | - | 4.0 (3.0, 6.0) | - | 6.0 (4.0, 10.0) |
| Total number of people over 18(IQR) | - | 6.0 (4.0, 9.0) | - | 2.0 (2.0, 3.0) | - | 3.0 (2.0, 5.0) |
| Total number of people under 18(IQR) | - | 6.0 (4.0, 10.0) | - | 2.0 (1.0, 3.0) | - | 3.0 (1.0, 5.0) |
| Father living with the child, N(%) | - | 701 (76.4) | - | 1708 (82.8) | - | 2409 (80.8) |
| Missing | - | 30 (3.3) | - | 50 (2.4) |  | 80 (2.7) |
| Mother was pregnant at the visit, N(%) | - | 75 (8.2) | - | 47 (2.3) | - | 122 (4.1) |

The PRECISE-DYAD study population was representative of the main PRECISE cohort in both countries, with participants demonstrating comparable demographic profiles (Table 1); 92.6% of the participants in Kenya and 97.5% in Gambia were reporting being married or cohabiting. In Kenya, approximately 92.5% of women had attained at least primary school education, whereas in The Gambia, 63.3% of women had no formal schooling, likely reflecting attendance at Koranic (Arabic) schools, which are not classified as formal education in international comparisons. More than half of participants across all sites were housewives, ranging from 52.9% in Kenya to 88% in The Gambia, and 34.4% of participants had returned to work or school by the time of the follow-up visit. Religious affiliation also differed, with the Kenyan cohort was more diverse (approximately 37.9% Muslim and 61.6% Christian) while Gambian women were almost exclusively Muslim (99.3%), and were representative of the main PRECISE cohort. Household composition varied between countries. In The Gambia, women reported living in larger households, with an average of six people under the age of 18 and six adults per household. In contrast, Kenyan households had a smaller average size, with two people under 18 and two adults. At the first visit the woman attended, 2.3% Kenyan participants and 8.2% Gambian participants reported being pregnant.

#### Maternal and Birth Outcomes

Analysis of pregnancy outcomes in the PRECISE-DYAD study population revealed differences compared with the primary PRECISE cohort (in this manuscript we have excluded the Mozambican data), with no evidence of enrichment in adverse birth outcomes overall, but a higher proportion of participants with hypertensive disorders. In the PRECISE-DYAD cohort, 28.9% of participants experienced hypertension during pregnancy, including 10% diagnosed with pre-eclampsia. This is similar as the prevalence of hypertensive disorders in the overall PRECISE pregnancy cohort (28.1% hypertensive women including 9,9% diagnosed with pre-eclampisa) (Table 2a).

**Table 2a.** Pregnancy outcomes of participants (from Kenya and The Gambia) originally recruited into the PRECISE study and subsequently followed up via the PRECISE-DYAD study. These tables describe pregnancy and birth outcomes among women retained in the PRECISE-DYAD follow-up is presented, alongside data from the original PRECISE cohort for comparison. Overall, there was no evidence of enrichment in adverse pregnancy and birth outcomes in the DYAD cohort; Data are shown to demonstrate the comparability between participants retained in postnatal follow-up and the overall pregnancy cohort. \**7 mothers unable to attend DYAD - child brought by caregiver* ICU – intensive care unit

|  | The Gambia |  | Kenya |  | Both countries |  |
| --- | --- | --- | --- | --- | --- | --- |
|  | PRECISE<br>Pregnancy | PRECISE<br>-DYAD<br>(first visit<br>attended) | PRECISE<br>Pregnancy | PRECISE<br>-DYAD<br>(first visit<br>attended) | PRECISE<br>Pregnancy | PRECISE<br>-DYAD<br>(first visit<br>attended) |
| **Number of women\*** | 1267 | 920 | 2769 | 2072 | 4036 | 2992 |
| **Maternal hypertension, N(%)** | 437 (34.5) | 345 (37.5) | 695 (25.1) | 520 (25.1) | 1132 (28.1) | 865 (28.9) |
| *Maternal gestational hypertension, N(%)* | 249 (19.7) | 206 (22.4) | 487 (17.6) | 362 (17.5) | 736 (18.2) | 568 (19.0) |
| *Maternal chronic hypertension, N(%)* | 188 (14.8) | 139 (15.1) | 204 (7.4) | 156 (7.5) | 392 (9.7) | 295 (9.9) |
| *Maternal preeclampsia, N(%)* | 172 (13.6) | 135 (14.7) | 228 (8.2) | 166 (8.0) | 400 (9.9) | 301 (10.1) |
| Missing maternal hypertension outcome, N(%) | 90 (7.1) | 21 (2.3) | 69 (2.5) | 50 (2.4) | 159 (3.9) | 71 (2.4) |
| **ICU admission, N(%)** | 2 (0.2) | 1 (0.1) | 0 (0.0) | 0 (0.0) | 2 (0.05) | 1 (0.0) |
| **Maternal death at birth** | 3 | 1 | 5 | 2 | 8 | 3 |

In PRECISE-DYAD, participants who experienced a miscarriage (0.3%), stillbirth (2.6%), or infant death (2.2%)(Table 2b), were invited to take part, and the representation of these groups was overall similar to that observed in the main PRECISE cohort (0.4 % miscarriage and 3.1% stillbirth). The proportion of twins (4.1%), was similar to the overall PRECISE cohort (4.2%). Overall, 36.2% children were classified as Small and Vulnerable Newborns (SVN). On the whole, 16.5% children were Small-for-Gestational Age (SGA; birthweight <10th percentile for sex and gestational age), 20.6% were born preterm (<37^+0^ weeks), 11.9% had low birthweight (LBW; birthweight <2500g). The prevalence of SGA, preterm and LBW amongst children recruited to DYAD mirrored that of the main PRECISE cohort; 1.9% of children had low (<7) Apgar scores measured 5 minutes after birth, with 56 children (1.8%), having a history of neonatal hospital admission.

**Table 2b.**
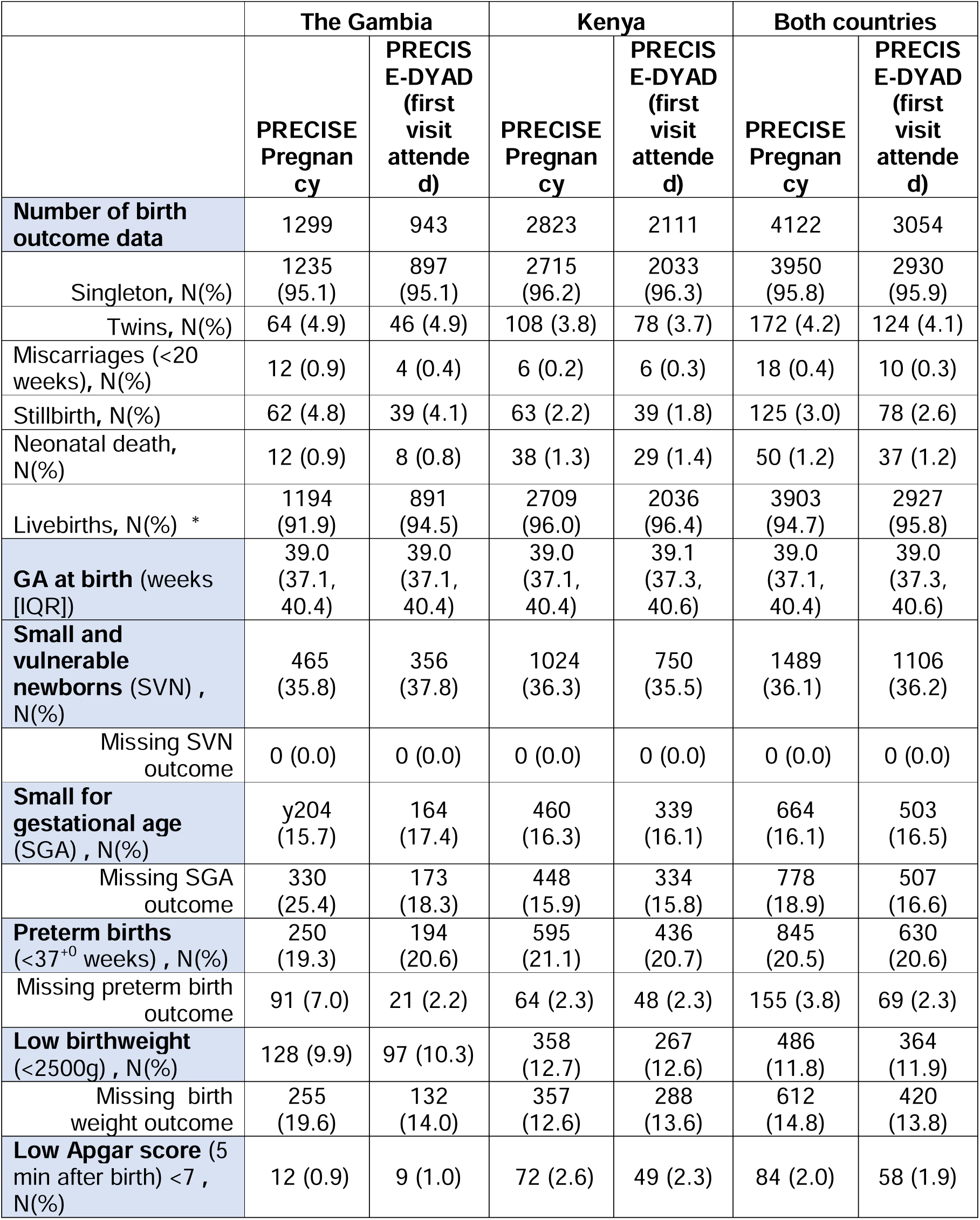
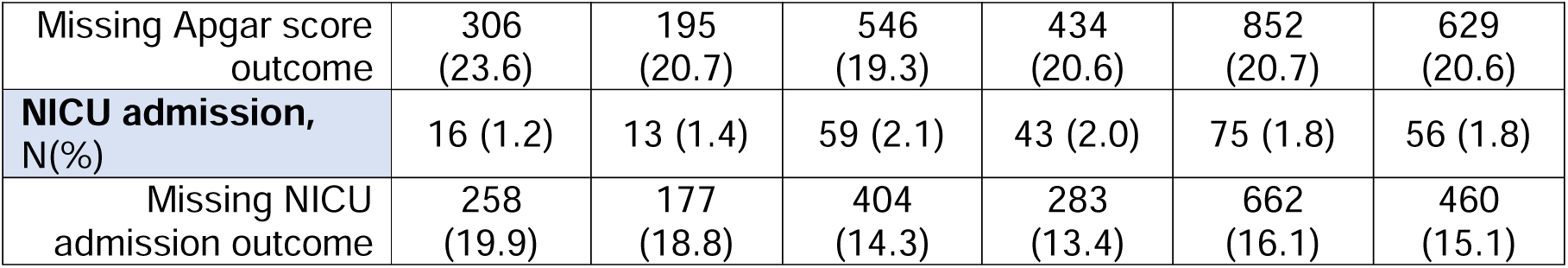
Birth outcomes of children (from Kenya and The Gambia) born during the PRECISE study and subsequently followed up via the PRECISE-DYAD study. IQR: Inter Quartile Range, NICU: neonatal intensive care unit, SGA: small for gestational age, SVN: small and vulnerable newborns. *2 children were reported dead in DYAD but are missing a date of death (unclassified): 1 was in Kenya and 1 in The Gambia

Mothers and infants were assessed either as dyads (mother-child) or individually; for participants seen without their biological mother or infant, the corresponding birth outcomes are presented in Table 2. Children whose mothers had died or were unable to participate to the study for unknow reasons were also followed up (Table 2a and b). Participants with pregnancy and birth outcome across visits are provided in Table S2.

#### Clinical Characteristics of Mother Participants

The maternal clinical characteristics are provided for the first visit the participant attended, which is 5.1 months after birth (IQR: 3.2, 12.4). Maternal nutritional status, as indicated by median Body Mass Index (BMI) was within the normal range (18.5-24.9kg/m^2^) for 53.6% of participants in both countries (Table 3a), the Gambian population tended to be more underweight (<18.5kg/m^2^, 18.4%), whereas the Kenyan population had a higher proportion of overweight women (>25kg/m^2^, 38.6%). In contrast, mid-upper arm circumference (MUAC), did not reflect these BMI categories, with 34.4% of women classified within the normal range and the prevalence of underweight and overweight women were similar in both countries. Minimum dietary diversity was met in 59.8% of participants.

**Table 3.** Maternal clinical characteristics at the first attended PRECISE-DYAD visit. This table presents maternal clinical data collected at the first postnatal visit attended by each participant. Measures include nutritional status (body mass index [BMI] and mid-upper arm circumference [MUAC]), cardiovascular assessments (including blood pressure), and mental health indicators. Data are shown to summarise the health profile of mothers at the start of the DYAD follow-up. IQR Inter Quartile Range, WHODAS: WHO Disability Assessment Schedule. ^#^122 women were pregnant at the first visit attended. * Full details of follow up by visit and timing presented in Table S2

|  | **The Gambia**<br>**PRECISE-DYAD**<br>**(first visit**<br>**attended)** | **Kenya**<br>**PRECISE-**<br>**DYAD (first**<br>**visit attended)** | **Both countries**<br>**PRECISE-DYAD**<br>**(first visit**<br>**attended)** |
| --- | --- | --- | --- |
| **Number of participants<sup>#</sup>**<br>**(total)** | 918 | 2034 | 2952 |
| **Interval of time between**<br>**birth and first visit attended**<br>**(months), median (IQR)** | 5.9 (5.4, 24.5) | 3.5 (3.0, 5.0) | 5.1 (3.2, 12.4) |
| **Maternal BMI, median (IQR)** | 21.6 (19.2, 25.0) | 23.6 (20.7,<br>27.5) | 23.1 (20.1, 26.7) |
| **Maternal BMI Category, N(%)** |  |  |  |
| Underweight <18.5 kg/m<sup>2</sup> | 169 (18.4) | 175 (8.6) | 344 (11.7) |
| Normal weight 18.5-24.9<br>kg/m<sup>2</sup> | 517 (56.3) | 1064 (52.3) | 1581 (53.6) |
| Overweight 25-29.9 kg/m<sup>2</sup> | 156 (17.0) | 493 (24.2) | 649 (22.0) |
| Obese 30 kg/m<sup>2</sup> | 68 (7.4) | 292 (14.4) | 360 (12.2) |
| Missing | 8 (0.9) | 10 (0.5) | 18 (0.6) |
| **MUAC- median (IQR)** | 27.1 (24.7, 30.0) | 27.1 (24.5,<br>30.2) | 27.1 (24.5, 30.2) |
| **MUAC Category, N(%)** |  |  |  |
| Underweight (<23.0cm) | 89 (9.7) | 218 (10.7) | 307 (10.4) |
| Normal weight (23.0-26.4cm) | 321 (35.0) | 693 (34.1) | 1014 (34.4) |
| Overweight (26.5 cm-29.9cm) | 272 (29.6) | 575 (28.3) | 847 (28.7) |
| Obese (≥30.0cm) | 231 (25.2) | 545 (26.8) | 776 (26.3) |
| Missing | 5 (0.5) | 3 (0.1) | 8 (0.3) |
| **Nutrition Status** |  |  |  |
| Meets minimum dietary<br>diversity, N(%) | 698 (76.0) | 1066 (52.4) | 1764 (59.8) |
| **Blood pressure Category,**<br>**N(%)** |  |  |  |
| Normal | 725 (78.9) | 1449 (71.2) | 2174 (73.6) |
| Elevated | 40 (4.4) | 115 (5.7) | 155 (5.3) |
| Stage 1 hypertension | 115 (12.5) | 370 (18.2) | 485 (16.4) |
| Stage 2 hypertension | 34 (3.7) | 98 (4.8) | 132 (4.5) |
| **Cardiology assessment**<br>**(IQR)** |  |  |  |
| Pulse wave velocity (m/sec) | 7.1 (6.5, 8.2) | 7.1 (6.6, 7.9) | 7.1 (6.5, 8.2) |
| Cardiac Output (L/min) | 4.2 (3.4, 4.9) | 4.8 (4.0, 5.8) | 4.2 (3.4, 4.9) |
| Systemic Vascular Resistance<br>(dynes.sec.cm-5) | 1578.9<br>(1355.8, 2020.9) | 1401.5<br>(1150.6, 1774.8) | 1578.9<br>(1355.8, 2020.9) |
| **Mental health , N(%)** |  |  |  |
| Number of participants who<br>screened positive for anxiety | 5/490 (1.0) | 22/662 (3.3) | 27/1152 (2.3) |
| Number of participants who<br>screened positive for<br>depression | 3/490 (0.6) | 12/662 (1.8) | 15/1152 (1.3) |
| Number of participants who<br>screened positive for post<br>traumatic stress | 2/365 (0.5) | 24/473 (5.1) | 26/838 (3.1) |
| Number of participants who<br>screened positive for<br>WHODAS | 1/489 (0.2) | 32/ 1680 (1.9) | 33/2169 (1.5) |
| Number of participants who<br>had suicidal thoughts | 6/487 (1.2) | 19/606 (3.1) | 25/1093 (2.3) |

Cardiovascular assessments were conducted one year postpartum using two devices. The arteriograph [22] [22], was used to measure arterial stiffness through pulse wave velocity. The Ultrasonic Cardiac Output Monitor (USCOM) [23] [23], was used to measure cardiac output (volume of blood the heart pumps per minute) and systemic vascular resistance (resistance that blood encounters as it flows through the blood vessels). All measurement fell between normal range, pulse wave velocity was at 7.1 m/s (normal range 6-9m/s), cardiac output 4.2 L/min (normal range 4–8 L/min) and systemic vascular resistance was 1578.9 dynes·s/cmD (normal range 700 to 1,600 dynes·s/cmD). At their first attended visit, 73.6% of women had a normal blood pressure and 20.9% have hypertension (systolic blood pressure ≥140mmHg or diastolic blood pressure ≥90mmHg). These data will contribute to understanding potential long-term cardiovascular risk factors among women in the cohort.

#### Mental Health

Mental health was assessed in a subset of participants using a panel of standardised tools. Initially this pilot study aimed at looking at the feasibility of asking women in these settings about their mental health [17] and was latter expanded to the full cohort at every visits to understand the influence of maternal health on neurodevelopment. Depression was measured with the Patient Health Questionnaire (PHQ-9) [24] [24], anxiety with the Generalised Anxiety Disorder scale (GAD-7) [25–27], post-traumatic stress disorder with the PTSD Checklist (civilian version (PLC-C) [28] [28], and the functional impairment with the World Health Organization Disability Assessment Scheduled (WHODAS 2.0) [29]. Overall, 2.3% of the participants screened positive for depression (PHQ-9), 1.3% for anxiety (GAD-7), 3.1% for PTSD (PCL-C), and 1.5% for functional impairment (WHODAS 2.0).

Additionally, 2.3% of the women reported experiencing suicidal thoughts. Despite a second training session in February 2023 in both countries that resulted in more women meeting the threshold in Kenya, the prevalence of anxiety and depression was slightly higher in Kenya compared with The Gambia. However, the overall prevalence rates remained lower than those reported in previous studies [30, 31]. All women who showed signs for depression, anxiety, or suicidal ideation were referred for follow-up through the existing clinical care pathways. Additional visit details are summarised in Table S2.

#### Clinical Characteristics of Child Participants

Child clinical data are presented based on the child latest visit, 23.1 months (IQR: 11.3, 35.0) in Table 3b. Females and males were equally represented in the follow-up, each accounting for approximately half of the participants (48.9% females and 50.5% males). Overall, 6.1% of the children required overnight hospital admission, with an average duration of 5.0 [3.0-7.3] days. The main reasons for hospitalisation were pneumonia, gastroenteritis, malnutrition, and infection. Overall, 42.2% of children were reported to have been tested for malaria. Of these tested, 13.3% of children were screened positive for malaria. Cough without fever or illness was reported in 16.7% children and 8.6% children had wheezing or whistling in the chest. Vision difficulty was reported in 0.3% of children, and 0.2% had hearing difficulties.

Exclusive breastfeeding was reported for 73.6% of children until six months of age (Table S3). Nutritional assessments indicated that 19.5% of the children were stunted, and 9% were wasted). MUAC measurement below −2 SD was found in 57.7% of children with 5.2% of the children who had a MUAC measurement <12.5cm, indicating moderate malnutrition. Blood pressure readings above 90^th^ percentile SD were found in 44.1% of children, while 4.1% were below the 10^th^ percentile using the pediatric blood pressure profile reference tool [32].

#### Neuro Development

Children were assessed at each visit for development using the Malawi Developmental Assessment Tool (MDAT) [33] which measures gross motor, fine motor, language and social domains as well as overall development with a Z score created through normative data from previous research in a number of different African countries (although mainly Malawi) https://kieran-bromley.shinyapps.io/mdat_scoring_shiny/. At their latest visit, 9.8% of the children fell below −1 SD for overall developmental and 2.5% below −2 SD (Table 3b). Starting at two years of age (visit 3), all children were screened using the Neurodevelopment Screen Tool (NDST) [34]. Of these children, 2.5% were at risk of developmental disabilities. Children at visit 3 were also screened for epilepsy, with 1.5% of children screening positive. For these children, the full history of epilepsy was administered to provide additional information of their previous epileptic seizure [35]. At the age of two, 76 children did not meet key developmental milestones (screened positive with MDAT or NDST) during the study and underwent additional assessments, a screen of visual acuity using the Cardiff cards, and the Modified Checklist for Autism in Toddlers (M-CHAT) to assess risk of autism spectrum disorder [36]. Of these, 24 children (30.7%) screened positive for autism and 67 (89.3%) screened positive for the visual acuity tests. Table S3 summarises the number of children who had the neurodevelopment assessment at each visit and by country.

#### Air Quality Sub-Study

A total of 343 women were recruited recruited from The Gambia, Kenya, and Mozambique. Among these participants, 76 (22.4%) women experienced hypertension during pregnancy, 38 (11.2%) were diagnosed with pre-eclampsia and 9 (2.7%) suffered a stillbirth. In this cohort, 133 (39.3%) children were classified as SVN, 77 (22.7%) were SGA (<10th percentile), 61 (18.0%) were born preterm, and 32 (9.5%) had low birth weight (Table 4). Exposure monitoring was conducted over 328 days between March 2022 and January 2023, covering both dry and wet seasons. This allowed the capture of a broad range of environmental conditions, settings, and pollution profiles. Overall, personal exposure to fine particulate matter (PMD.D) was 30.8 µg/m³ (IQR: 12.3–37.6 µg/m³), exceeding the World Health Organization (WHO) air quality guidelines (5 µg/m³ for annual exposure and 15 µg/m³ for short- to long-term exposure) across all sites (WHO 2021). Peak personal exposure reached 491.6 µg/m³ (IQR: 154.9–1052.4 µg/m³).Second PRECISE Pregnancies

**Table 4.** Children’s clinical characteristics at the latest PRECISE-DYAD visit. This table presents clinical data for children based on their most recent postnatal visit, including hospital admissions, malaria testing, history of cough, hearing or vision difficulties, and nutritional status (mid-upper arm circumference [MUAC]). Data are shown to summarise the health profile of children at their latest follow-up. BP: blood pressure: MDAT: Malawi Development Assessment Tool, NDST: neurodevelopmental screening tool, IQR Inter Quartile Range, MUAC: mid-upper arm circumference, SD Standard deviation

|  | **The Gambia**<br>**PRECISE-DYAD**<br>**(latest visit**<br>**attended)** | **Kenya**<br>**PRECISE-DYAD**<br>**(latest visit**<br>**attended)** | **Both countries**<br>**PRECISE-DYAD**<br>**(latest visit**<br>**attended)** |
| --- | --- | --- | --- |
| **Number of participants** | 883 | 1998 | 2881 |
| Age(months) - Median (IQR) | 24.8 (23.7, 35.8) | 23.0 (11.0, 23.6) | 23.1 (11.3, 35.0) |
| Girls, N(%) | 436 (49.4) | 972 (48.6) | 1408 (48.9) |
| Boys, N(%) | 445 (50.4) | 1013 (50.7) | 1458 (50.6) |
| Missing | 2 (0.2) | 13 (0.7) | 15 (0.5) |
| **Child health, N(%)** |  |  |  |
| Hospital admission | 25 (2.8) | 150 (7.5) | 175 (6.1) |
| missing | 14 (1.6) | 9 (0.5) | 23 (0.8) |
| Hospital stay length- days-<br>median (IQR) | 7.0 (3.0, 14.0) | 5.0 (3.0, 7.0) | 5.0 (3.0, 7.3) |
| Malaria test | 91 (10.3) | 1124 (56.3) | 1215 (42.2) |
| Missing malaria test | 11 (1.2) | 6 (0.3) | 17 (0.6) |
| Test result positive | 2 (2.2) | 158 (14.1) | 160 (13.3) |
| Missing test result | 0 (0.0) | 0 (0.0) | 0 (0.0) |
| Child cough when no fever or<br>illness | 73 (8.3) | 408 (20.4) | 481 (16.7) |
| Missing | 48 (5.4) | 247 (12.4) | 295 (10.2) |
| Child has wheezing or<br>whistling in the chest | 26 (2.9) | 223 (11.2) | 249 (8.6) |
| Missing | 47 (5.3) | 245 (12.3) | 292 (10.1) |
| Child reported with vision<br>difficulty | 1 (0.1) | 8 (0.4) | 9 (0.3) |
| Child reported with hearing<br>difficulty | 2 (0.2) | 5 (0.3) | 7 (0.2) |
| Missing seeing/hearing<br>difficulty | 46 (5.2) | 245 (12.3) | 291 (10.1) |
| **Child nutrition Status** |  |  |  |
| **Stunting z-score** | -1.3 (-1.9, -0.6) | -0.9 (-1.8, -0.1) | -1.1 (-1.8, -0.2) |
| Number of Children Stunted,<br>N(%) | 176 (19.9) | 387 (19.4) | 563 (19.5) |
| **Wasting z-score** | -0.8 (-1.5, -0.2) | -0.3 (-1.1, 0.5) | -0.5 (-1.2, 0.3) |
| Number of Children Wasted,<br>N(%) | 119 (13.5) | 141 (7.1) | 260 (9.0) |
| Missing stunting and wasting | 0 (0.0) | 0 (0.0) | 0 (0.0) |
| **MUAC Z score** | -0.94 (-1.55, - | -0.21 (-0.92, 0.49) | -0.48 (-1.14, 0.28) |
|  | 0.35) |  |  |
| MUAC under threshold <-2SD, N(%) | 633 (71.7) | 1030 (51.5) | 1663 (57.7) |
| Number of children with MUAC average <11.5 cm (severe malnutrition) , N(%) | 11 (1.2) | 26 (1.3) | 37 (1.3) |
| Number of children with MUAC average ≥11.5, <12.5cm (moderate malnutrition) , N(%) | 46 (5.2) | 66 (3.3) | 112 (3.9) |
| Missing | 10 (1.1) | 6 (0.3) | 16 (0.6) |
| **Child Blood pressure, N(%)** |  |  |  |
| Number of children with BP ≥90th percentile | 519 (58.7) | 765 (38.3) | 1284 (44.6) |
| Number of children with BP <10th percentile | 3 (0.3) | 114 (5.7) | 117 (4.1) |
| **Neuro Assessment, N(%)** |  |  |  |
| Number of children screened positive MDAT <-1SD | 104 (11.8) | 177 (8.9) | 281 (9.8) |
| Number of children screened positive MDAT <-2SD | 19 (2.2) | 54 (2.7) | 73 (2.5) |
| Number of children at risk of developmental delay (NDST) | 8 (0.9) | 65 (3.3) | 73 (2.5) |
| Number of children screened positive epilepsy | 2 (0.2) | 40 (2.0) | 42 (1.5) |
| Number of children screened positive for MCHAT | 9/16 (56.3) | 15/60 (23.7) | 24/76 (30.7) |
| Number of children screened positive for CARDIFF | 15/16 (93.8) | 52/60 (88.1) | 67/76 (89.3) |

**Table 5.** Pregnancy outcomes of participants recruited in the air quality substudy. This table summarises the number of participants included in the air quality sub study, their pregnancy outcomes, and measured personal exposure to particulate matter (PMD.D) using portable sensor packs. Data are presented to illustrate individual exposure levels and to contextualise outcomes within the sub study population.

| **Air quality** | **The Gambia** | **Kenya** | **Mozambique** | **All countries** |
| --- | --- | --- | --- | --- |
| **Number of participants** | 160 | 105 | 78 | 343 |
| **pregnancy outcome, N(%)** |  |  |  |  |
| gestational hypertension | 41 (25.6) | 27 (25.7) | 8 (10.3) | 76 (22.2) |
| Pre-eclampsia | 22 (14.4) | 15 (14.3) | 1 (1.3) | 38 (11.4) |
| missing birth outcome | 0 | 0 | 5 | 5 |
| **Number of children** | 164 | 105 | 74 | 343 |
| Low Birth Weight, N(%) | 20 (12.2) | 6 (5.7) | 6 (8.1) | 32 (9.3) |
| Large for gestational age >95th percentile, N(%) | 9 (5.5) | 13 (12.4) | 5 (6.8) | 27 (7.9) |
| Small for gestational age <3rd percentile, N(%) | 47 (28.7) | 14 (13.3) | 16 (21.6) | 77 (22.4) |
| Preterm <37 weeks, N(%) | 27 (16.5) | 24 (22.9) | 10 (13.5) | 61 (17.8) |
| Small vulnerable newborn, N(%) | 70 (42.7) | 37 (35.2) | 26 (35.2) | 133 (38.8) |
| Stillbirth, N(%) | 8 (4.9) | 0 | 1 (1.3) | 9 (2.6) |
| **Interval between delivery to first PM<sub>2.5</sub> assessment, months** | 11 (5-12) | 5 (4-10) | 9 (7-11) | 9 (5-12) |
| Monitoring period start, YYYY-MM-DD | 2022-03-31 | 2022-03-09 | 2022-05-04 | 2022-03-09 |
| Monitoring period end, YYYY-MM-DD | 2023-01-31 | 2022-12-13 | 2022-10-11 | 2023-01-31 |
| **Personal PM<sub>2.5</sub> exposure** |  |  |  |  |
| Mean daily exposure (IQR) | 31.6<br>(12.0-39.7) | 32.<br>(12.9-27.8) | 24.3<br>(12.0-27.8) | 30.8<br>(12.3-37.6) |
| Peak daily exposure(IQR) | 489.1<br>(155.2-1052.4) | 842.0<br>(263.8-1052.4) | 172.2<br>(75.3-482.4) | 491.6<br>(154.9-1052.4) |

**Table 6.**
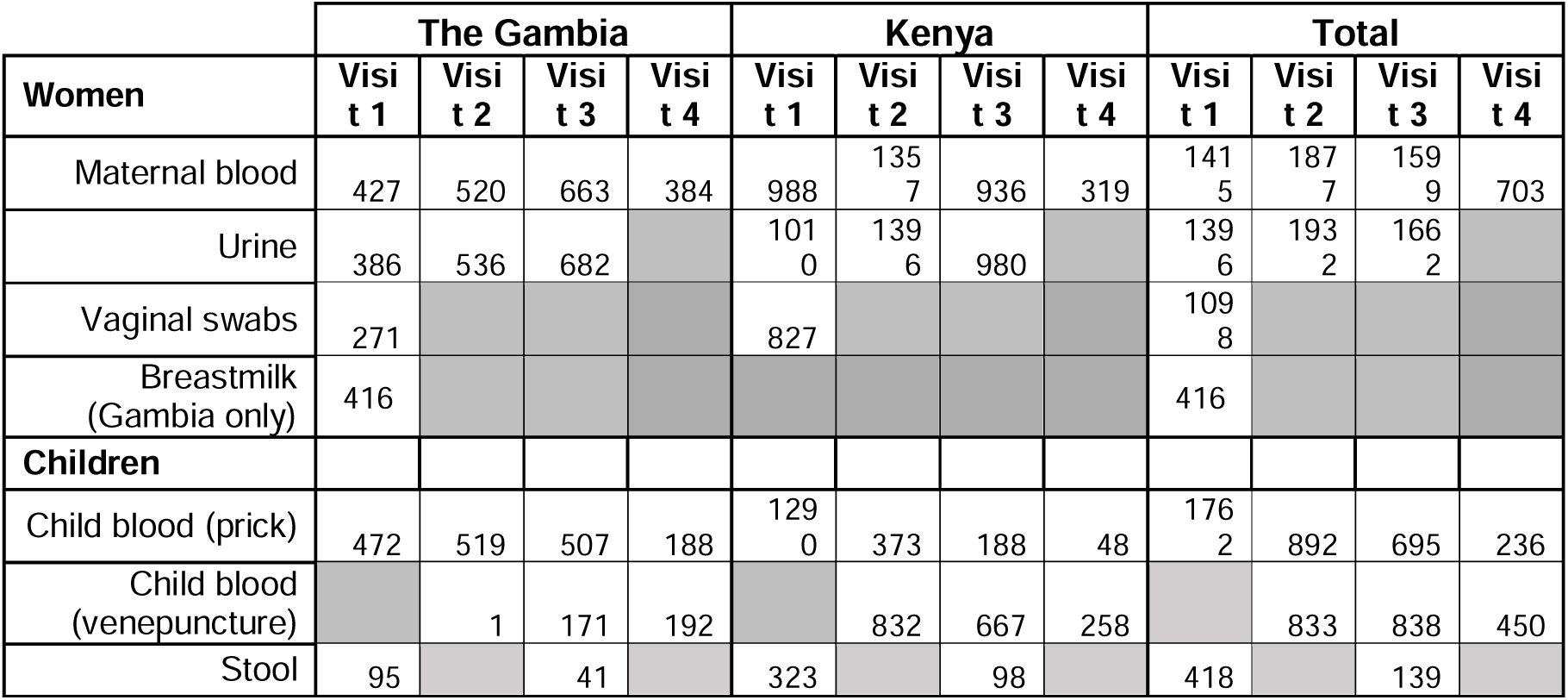
Number of women and children with samples collected during PRECISE-DYAD study. This table summarises the number of women and children from whom biological samples were successfully collected at each PRECISE-DYAD study visit. Maternal samples include blood, urine, vaginal swabs, and breastmilk (The Gambia only). Children’s samples include blood collected via finger prick or venepuncture, and stool. Data are shown to illustrate sample availability across the follow-up period.

#### PRECISE in DYAD Sub-Study

During the PRECISE and PRECISE-DYAD studies, 141 women who became pregnant during the study were enrolled in a shorter version of the PRECISE, as described in Craik *et al.* (2022) [17]. These data provide insight into how previous pregnancy complications may influence decisions around birth spacing, as well as the potential impact of raising a child with moderate-to-severe neurodevelopmental disability (versus those without such outcomes) on subsequent pregnancies. In some cases, women were re-recruited into the study following an earlier miscarriage or pregnancy loss.

### Biological Samples

The PRECISE-DYAD study established an extensive biorepository of samples collected from both women and children across multiple study visits. For women, samples included maternal blood (blood spots, buffy coat, plasma, and serum), urine, vaginal swabs and breastmilk (collected in The Gambia only). For children, samples included blood (via heel prick or venepuncture) and stool specimens. The number of women and children from whom samples were collected at each study visit is shown in Table 4. A detailed breakdown of the samples collected can be found in Table S5.

#### Forthcoming analyses

Several analyses using the PRECISE-DYAD sample and data are currently underway. These include evaluations of the impact of pregnancy complications on child neurodevelopmental outcomes and on maternal cardiovascular health one year postpartum. Additional ongoing work is assessing associations between personal exposure to air pollutants and heat stress during pregnancy and adverse maternal, perinatal, infant, and neurodevelopmental outcomes. The effects of SARS-CoV-2 infection in pregnancy on child neurodevelopment are being investigated, alongside efforts to develop methodological approaches for assessing mental health within large longitudinal studies.

A health economic analysis is examining how pregnancy complications affect household economic resilience, including direct medical costs, income loss, and food security. This nested case–control sub-study examined the financial burden associated with pregnancy and childbirth, particularly in the context of complications. A total of 210 participants were recruited and interviewed at their home, the details of the case–control distribution is summarised in Table S4.

The quality of maternal and newborn care sub-study explored women’s experiences of the care they and their newborn, received during labour and birth within healthcare facilities, and if these differed for women who experienced an adverse pregnancy outcome compared with those with uncomplicated pregnancies. A total of 1,132 participants were recruited to this sub-study; the distribution of the case-control cohort is summarised in table S4 (Table S4).

A total of 457 water samples were collected 222 from community water sources (97 in Kenya and 125 in The Gambia) and 235 from participants’ households (99 in Kenya and 136 in The Gambia). Water samples are being tested to assess seasonal variation and cross-country differences, contributing to understanding environmental risks associated with water quality for maternal and child health Microbiological testing was conducted on all samples to detect bacterial contamination, specifically total coliforms and *Escherichia coli*. Additionally, source water samples were tested for physico-chemical parameters (including pH, total dissolved solids, conductivity, residual chlorine, and major ions) and heavy metals (lead, arsenic, and mercury).

An initial quality control assessment was conducted on a subset of biological samples to evaluate their suitability for downstream genetic analysis. Twenty buffy coat samples from each site were assessed, alongside twenty stool swabs from Kenya. Samples were selected based on processing time (i.e., the time elapsed from collection to freezing), including the five fastest and five slowest processed samples per type and site. DNA was extracted from each sample, quantified, and assessed using gel electrophoresis to evaluate yield, purity (A260/A280 ratio), and evidence of degradation. Overall, the samples demonstrated acceptable DNA quantity, purity, and integrity for use in genetic analysis. Finally, stool samples from children are undergoing analysis to characterise the gut microbiota and explore its associations with child health outcomes.

### Publications to Date

The methodology for collecting clinical and biological data in the PRECISE-DYAD study has been published [17], alongside a detailed description of the neurodevelopmental assessment methods [18]. A systematic review of respectful maternity care training packages for health workers in sub-Saharan Africa has also been published [37] [37]. Moreover, we have evaluated the association between household water, sanitation, and hygiene (WASH) status and pregnancy complications across The Gambia, Kenya, and Mozambique [12].

## Discussion

This study provides a uniquely rich, pregnancy-enrolled, population-based cohort that combines extensive social, clinical, and biological data—including biospecimens—from two geographically and culturally distinct settings in sub-Saharan Africa. Recruiting women during pregnancy enabled early identification and longitudinal follow-up of those with placenta-related complications. By integrating data from both PRECISE and PRECISE-DYAD, the study offers an unparalleled opportunity to examine the determinants and consequences of placental disorders on both maternal and child health trajectories (and the interaction between those trajectories), while generating additional and valuable insights into outcomes influenced by the COVID-19 pandemic.

A limitation is the loss to follow-up from enrolment during pregnancy to up to 3 years after birth. Overall, of the 4,122 women who had a PRECISE birth visit, 2,980 attended at least one DYAD visit, giving a follow-up rate of 72.3% overall. When calculated against the full PRECISE enrolment cohort of 5,026 women, the overall follow-up rate fell to 59.3%, with 56.2% in Kenya and 67.7% in The Gambia. Tracing was more effective in The Gambia, where most women were part of the Health and Demographic Surveillance Systems (HDSS). In Kenya, however, only 60% of rural and 30% of urban women resided within the HDSS area, making follow-up significantly more challenging despite substantial effort and resource investment. Contributing factors to loss to follow-up included the extended time between initial recruitment and eligibility for PRECISE-DYAD follow-up (e.g., women who gave birth in 2019 only became eligible to the study in 2022), relocation, and changes in contact information. This loss to follow-up may have been further exacerbated by the COVID-19 pandemic that happened during the PRECISE study and disrupted routine health services and reduced participants’ ability to attend visits. These challenges highlight the critical importance of sustained engagement with families in longitudinal studies. Our extensive community engagement, including 108 sensitisation meetings with nearly 4,000 women, likely played a key role in retaining these participants in the study despite the challenges of long-term follow-up.

### Defninitions

<u>Stunting</u> is defined as height-for-age more than two standard deviations below the WHO Child Growth Standards median

<u>Wasting</u> is defined weight-for-height more than 2 standard deviations below the WHO Child Growth Standards median.

<u>Child blood pressure</u> was calculated using pedbp package in R [32]

<u>Hypertension in pregnancy</u>: This is defined as a clinic systolic BP ≥ 140 mmHg and/or a diastolic BP ≥ 90 mmHg, with systolic BP ≥ 160 mmHg and/or a diastolic BP ≥ 110 mmHg defined as severe hypertension [38].

<u>Gestational hypertension</u>: This is defined as hypertension arising *de novo* at ≥ 20 weeks’ gestation in the absence of proteinuria or other findings suggestive of pre-eclampsia [38]

<u>Pre-eclampsia (*de novo*)</u>: This is defined as gestational hypertension accompanied by one or more of the following new-onset conditions at ≥ 20 weeks’ gestation:

- i) Proteinuria
- ii) Other maternal end-organ dysfunction, including neurological complications (e.g., eclampsia, altered mental status, blindness, stroke, clonus, severe headache, or persistent visual scotomata), pulmonary oedema, haematological complications (e.g., platelets <150,000/microlitre, disseminated intravascular coagulation, haemolysis), acute kidney injury (e.g., creatinine >90 µmol/litre or >1mg/dL), liver involvement (e.g., elevated transaminases with or without right upper quadrant or epigastric abdominal pain)
- iii) Uteroplacental dysfunction (e.g., placental abruptio, angiogenic imbalance foetal growth restriction or intrauterine fetal death) [38]

<u>Stillbirth</u>: This is defined as an infant born with no signs of life after a given threshold, usually related to the gestational age or weight of the baby; in this study we will use both the current World Health Organization (WHO) definition for international comparison of a stillbirth as being ‘a baby born without signs of life at or after 28 weeks of gestation [39], and the more inclusive definition of birth of an infant without signs of life ≥500g or ≥20^0^ weeks of gestation[40]).

<u>Preterm birth</u>: The WHO defines preterm birth as any birth before 37 completed weeks of gestation (fewer than 259 days since the first day of the women’s last menstrual period) [41].

<u>Small for gestational age</u>: This is defined as infants (ex utero) weighing less than the 10^th^ centile birth weight for gestational age and sex. We will use the multi-ethnic, INTERGROWTH-21^st^ birth weight standard [42, 43].

<u>Neonatal deaths</u>: The death of a live-born infant within the first 28 days of life [44]

<u>Small and Vulnerable Newborn</u> was defined as an infant born either preterm (<37 weeks 0 days gestation) or below the third percentile for sex and gestational age (Intergrowth-21^st^ chart) [45]

## Further Details

### Contributorship Statement

PVD, MT, AR, UDA, GO, BB, MG, AA, RT, AsK, TM, TTS, SM, HN, VF, LAM, LP, ES, HB, MAM were involved in the conception and design of the study. MLV, RC, AK, HJ, MoM, OW, IM, JM, FT, EM, YI, FK, KB, BN, MO, GM, JL, DM, JC, HDM, LM, AM, AR managed data collection. MLV, RO, MW, JA had significant input into the data cleaning and analysis. MLV drafted the final manuscript. All authors had significant input into the manuscript.

### Collaboration

We have built the combined PRECISE and PRECISE-DYAD biorepository to help strengthen research capacity in sub-Saharan Africa, whilst simultaneously investigating the impact of placental conditions in these communities. Our priority is to support researchers in Africa to utilise these data and samples to enable them to build their own careers and local research capacity. As part of this, each site team owns its own data and samples. Anyone is welcome to request the data; however, our preferred method of data sharing is to form collaborations with external groups to enable those who collected the data to be involved in its analysis. We believe that this strengthens the analysis, as the site teams best understand the complexities of their data, thereby enabling researchers to optimise the use and understand the implications of the data. To access data, please; you will be asked to complete a Research Application Form, outlining your research plans, which will be reviewed for scientific merit by our Data and Sample Access Committee.

### Data Availability Statement

Data are available on reasonable request. The data that support the findings of this study are available from the corresponding author on reasonable request.

### Ethics Approvals

Approval for the PRECISE-DYAD study was obtained from King’s College London (Ref HR-20/21-19714), The Gambia Government/MRC Joint Ethics Committee (Ref 22843), Aga Khan University, Nairobi Institutional Ethics Review Committee (Ref 2021/IERC-08) and University of British Columbia (Ref H20-02769). Approval for the PRECISE-HOME study was obtained in King’s College London (Ref HR-17/18–7855), the Mozambique Ministry of Health, National Bioethics Committee for Health (CIBS-CISM/105/2021). Approval for the PRECISE study was obtained in King’s College London (Ref HR-17/18–7855), Aga Khan University Hospital (Ref 2018/REC-74), The Gambia Government/The Medical Research Council, The Gambia Joint Committee (Ref SCC 1619), and the Mozambique Ministry of Health, National Bioethics Committee for Health (545/CNBS/18).

## Acknowledgments

We are extremely grateful to all the women and families who took part in this study, the hospital staff for their help in conducting the study, and the whole PRECISE team, which includes interviewers, fieldworkers, enumerators, laboratory technicians, clerical workers, research scientists, volunteers, managers, receptionists and nurses. In addition, we thank the PRECISE partner institutions and governments for their support.

