## Supplementary material for "Cohort Profile: PRECISE-DYAD: a prospective cohort study linking maternal and infant health trajectories in sub-Saharan Africa": Table s1

**Table S1** **Pregnancy and birth outcomes of participants (from Kenya and The Gambia) at each PRECISE-DYAD visits.**

|  | **The Gambia** | | | | **Kenya** | | | | **All countries** | | | |
| --- | --- | --- | --- | --- | --- | --- | --- | --- | --- | --- | --- | --- |
|  | Visit 1  (6 wk-6 months  after birth) | Visit 2  (12 months  after birth) | Visit 3 (24 months after  birth) | Visit 4 (36 months  after birth) | Visit 1  (6 wk-6 months  after birth) | Visit 2  (12 months  after birth) | Visit 3 (24 months after  birth) | Visit 4 (36 months  after birth) | Visit 1  (6 wk-6 months  after birth) | Visit 2  (12 months  after birth) | Visit 3 (24 months after  birth) | Visit 4 (36 months  after birth) |
| Number of women recruited in PRECISE-DYAD | **440** | **542** | **727** | **420** | **1526** | **1483** | **1109** | **402** | **1966** | **2025** | **1836** | **822** |
| Miscarriages (<20 weeks) N (%) | 0 (0.0) | 2 (0.4) | 2 (0.3) | 0 (0.0) | 0 (0.0) | 5 (0.3) | 4 (0.4) | 2 (0.5) | 0 (0.0) | 7 (0.3) | 6 (0.3) | 2 (0.2) |
| Maternal hypertension (HDP) N (%) | 180 (40.9) | 231 (42.6) | 282 (38.8) | 139 (33.1) | 406 (26.6) | 395 (26.6) | 297 (26.8) | 108 (26.9) | 586 (29.8) | 626 (30.9) | 579 (31.5) | 247 (30.1) |
| *Maternal gestational hypertension* | 116 (26.4) | 136 (25.1) | 168 (23.1) | 80 (19.0) | 297 (19.5) | 272 (18.3) | 203 (18.3) | 65 (16.2) | 413 (21.0) | 408 (20.1) | 371 (20.2) | 145 (17.6) |
| *Maternal chronic hypertension* | 64 (14.5) | 95 (17.5) | 114 (15.7) | 59 (14.0) | 108 (7.1) | 122 (8.2) | 93 (8.4) | 43 (10.7) | 172 (8.7) | 217 (10.7) | 207 (11.3) | 102 (12.4) |
| *Maternal preeclampsia* | 75 (17.0) | 92 (17.0) | 110 (15.1) | 55 (13.1) | 133 (8.7) | 134 (9.0) | 95 (8.6) | 39 (9.7) | 208 (10.6) | 226 (11.2) | 205 (11.2) | 94 (11.4) |
| Missing maternal hypertension outcome N (%) | 21 (4.8) | 2 (0.4) | 1 (0.1) | 0 (0.0) | 49 (3.2) | 2 (0.1) | 1 (0.1) | 1 (0.2) | 70 (3.6) | 4 (0.2) | 2 (0.1) | 1 (0.1) |
| Maternal death at birth N (%) | 1 (0.2) | 1 (0.2) | 1 (0.1) | 0 (0.0) | 1 (0.1) | 1 (0.1) | 2 (0.2) | 0 (0.0) | 2 (0.1) | 2 (0.1) | 3 (0.2) | 0 (0.0) |
| ICU admission N (%) | 0 (0.0) | 0 (0.0) | 1 (0.1) | 1 (0.2) | 0 (0.0) | 0 (0.0) | 0 (0.0) | 0 (0.0) | 0 (0.0) | 0 (0.0) | 1 (0.1) | 1 (0.1) |
| Number of children recruited in PRECISE-DYAD | **452)** | **559** | **746** | **427** | **1554** | **1515** | **1132** | **410)** | **2006** | **2074** | **1878** | **837** |
| GA at birth (weeks [IQR]) | 39.0 (37.1, 40.6) | 39.0 (37.1,40.4) | 39.1 (37.3, 40.6) | 39.0 (37.3, 40.1) | 39.1 (37.3, 40.6) | 39.1 (37.2, 40.6) | 39.0 (37.0, 40.6) | 39.1 (37.6, 40.5) | 39.1 (37.1, 40.6) | 39.1 (37.1, 40.6) | 39.0 (37.1, 40.6) | 39.0 (37.6, 40.3) |
| Stillbirths N (%) | 16 (3.5) | 22 (3.9) | 29 (3.9) | 13 (3) | 22 (1.4) | 27 (1.8) | 21 (1.9) | 6 (1.5) | 38 (1.9) | 49 (2.4) | 50 (2.7) | 19 (2.3) |
| Infant death N (%) | 6 (1.3) | 8 (1.4) | 17 (2.3) | 9 (2.1) | 24 (1.5) | 32 (2.1) | 31 (2.7) | 11 (2.7) | 30 (1.5) | 40 (1.9) | 48 (2.6) | 20 (2.4) |
| Livebirths N (%) | 436 (96.5) | 535 (95.7) | 714 (95.7) | 413 (96.7) | 1532 (98.6) | 1482 (97.8) | 1107 (97.8) | 402 (98) | 1968 (98.1) | 2017 (97.3) | 1821 (97) | 815 (97.4) |
| Singleton N (%) | 428 (94.7) | 525 (93.9) | 708 (94.9) | 413 (96.7) | 1498 (96.4) | 1451 (95.8) | 1086 (95.9) | 394 (96.1) | 1926 (96) | 1976 (95.3) | 1794 (95.5) | 807 (96.4) |
| Twins N (%) | 24 (5.3) | 34 (6.1) | 38 (5.1) | 14 (3.3) | 56 (3.6) | 64 (4.2) | 46 (4.1) | 16 (3.9) | 80 (4) | 98 (4.7) | 84 (4.5) | 30 (3.6) |
| Small and vulnerable newborns (SVN) N (%) | 191 (42.3) | 229 (41) | 279 (37.4) | 147 (34.4) | 565 (36.4) | 546 (36) | 423 (37.4) | 123 (30) | 756 (37.7) | 775 (37.4) | 702 (37.4) | 270 (32.3) |
| Small for gestational age (SGA) N (%) | 99 (21.9) | 114 (20.4) | 137 (18.4) | 65 (15.2) | 254 (16.3) | 245 (16.2) | 188 (16.6) | 57 (13.9) | 353 (17.6) | 359 (17.3) | 325 (17.3) | 122 (14.6) |
| Missing SGA outcome | 79 (17.5) | 98 (17.5) | 136 (18.2) | 72 (16.9) | 181 (11.6) | 222 (14.7) | 182 (16.1) | 78 (19) | 260 (13) | 320 (15.4) | 318 (16.9) | 150 (17.9) |
| Preterm births N (%) | 89 (19.7) | 121 (21.6) | 150 (20.1) | 86 (20.1) | 325 (20.9) | 325 (21.5) | 255 (22.5) | 70 (17.1) | 414 (20.6) | 446 (21.5) | 405 (21.6) | 156 (18.6) |
| Missing preterm birth outcome | 21 (4.6) | 2 (0.4) | 1 (0.1) | 0 (0) | 48 (3.1) | 1 (0.1) | 0 (0) | 0 (0) | 69 (3.4) | 3 (0.1) | 1 (0.1) | 0 (0) |
| Low birthweight N (%) | 57 (12.6) | 72 (12.9) | 80 (10.7) | 36 (8.4) | 202 (13) | 199 (13.1) | 145 (12.8) | 46 (11.2) | 259 (12.9) | 271 (13.1) | 225 (12) | 82 (9.8) |
| Missing low birth weight outcome | 55 (12.2) | 74 (13.2) | 108 (14.5) | 56 (13.1) | 150 (9.7) | 189 (12.5) | 157 (13.9) | 70 (17.1) | 205 (10.2) | 263 (12.7) | 265 (14.1) | 126 (15.1) |
| Low Apgar score N (%) | 1 (0.2) | 3 (0.5) | 6 (0.8) | 7 (1.6) | 44 (2.8) | 39 (2.6) | 38 (3.4) | 6 (1.5) | 45 (2.2) | 42 (2) | 44 (2.3) | 13 (1.6) |
| Missing Apgar score outcome | 50 (11.1) | 88 (15.7) | 156 (20.9) | 129 (30.2) | 213 (13.7) | 276 (18.2) | 240 (21.2) | 129 (31.5) | 263 (13.1) | 364 (17.6) | 396 (21.1) | 258 (30.8) |
| NICU admission N (%) | 10 (2.2) | 11 (2) | 12 (1.6) | 3 (0.7) | 35 (2.3) | 30 (2) | 29 (2.6) | 6 (1.5) | 45 (2.2) | 41 (2) | 41 (2.2) | 9 (1.1) |
| Missing NICU admission outcome | 39 (8.6) | 71 (12.7) | 148 (19.8) | 121 (28.3) | 124 (8) | 157 (10.4) | 162 (14.3) | 100 (24.4) | 163 (8.1) | 228 (11) | 310 (16.5) | 221 (26.4) |

**Table S2** **Maternal demographic information and clinical characteristics of participants at each PRECISE-DYAD visits.**

|  | **The Gambia** | | | | **Kenya** | | | | **All countries** | | | |
| --- | --- | --- | --- | --- | --- | --- | --- | --- | --- | --- | --- | --- |
|  | Visit 1  (6 wk-6 months  after birth) | Visit 2  (12 months  after birth) | Visit 3  (24 months after  birth) | Visit 4 (36 months  after birth) | Visit 1  (6 wk-6 months  after birth) | Visit 2  (12 months  after birth) | Visit 3 (24 months after  birth) | Visit 4 (36 months  after birth) | Visit 1  (6 wk-6 months  after birth) | Visit 2  (12 months  after birth) | Visit 3 (24 months after  birth) | Visit 4 (36 months  after birth) |
| **Number of participants (total)** | 439 | 540 | 719 | 418 | 1523 | 1476 | 1079 | 380 | 1962 | 2016 | 1798 | 798 |
| Number of participants (in person) | 439 | 540 | 719 | 418 | 1515 | 1458 | 1053 | 363 | 1954 | 1998 | 1772 | 781 |
| Number of phone interview | 0 | 0 | 0 | 0 | 8 | 18 | 27 | 17 | 8 | 18 | 27 | 17 |
| **Interval of time between birth and visit (months), median (IQR)** | 5.5 (5.0, 5.8) | 11.8 (11.1, 12.6) | 24.0 (23.5, 24.6) | 35.9 (35.4, 36.4) | 3.6, 3.3 (3.0, 4.1) | 11.4, 11.1 (11.0, 11.7) | 23.1 (23.0, 23.6) | 35.1 (35.0, 35.5) | 3.5 (3.1, 5.0) | 11.2 (11.0, 12.0) | 23.4 (23.0, 24.2) | 35.5 (35.1, 36.2) |
| **Maternal age, years median (IQR)** | 28 (23, 32) | 28 (23, 32) | 29 (24, 33) | 30 (25, 34) | 27 (23, 32) | 28 (24, 33) | 30 (26, 34) | 31 (27, 36) | 27 (23, 32) | 28 (24, 33) | 29 (25, 34) | 30 (26, 35) |
| **Age category, years** N (%) |  |  |  |  |  |  |  |  |  |  |  |  |
| 15-19 | 26 (5.9) | 31 (5.7) | 14 (1.9) | 2 (0.5) | 54 (3.5) | 38 (2.6) | 4 (0.4) | 1 (0.3) | 80 (4.1) | 69 (3.4) | 18 (1.0) | 3 (0.4) |
| 20-24 | 113 (25.7) | 131 (24.2) | 167 (23.2) | 84 (20.1) | 425 (27.9) | 358 (24.3) | 177 (16.4) | 43 (11.3) | 538 (27.4) | 489 (24.3) | 344 (19.1) | 127 (15.9) |
| 25-29 | 130 (29.6) | 155 (28.7) | 203 (28.2) | 118 (28.2) | 459 (30.1) | 446 (30.2) | 345 (31.9) | 126 (33.2) | 589 (30.0) | 601 (29.8) | 548 (30.5) | 244 (30.6) |
| 30-34 | 92 (21.0) | 120 (22.2) | 182 (25.3) | 115 (27.5) | 342 (22.5) | 355 (24.1) | 293 (27.1) | 91 (23.9) | 434 (22.1) | 475 (23.6) | 475 (26.4) | 206 (25.8) |
| 35-39 | 50 (11.4) | 69 (12.8) | 100 (13.9) | 61 (14.6) | 191 (12.5) | 221 (15.0) | 197 (18.2) | 84 (22.1) | 241 (12.3) | 290 (14.4) | 297 (16.5) | 145 (18.2) |
| 40-44 | 25 (5.7) | 30 (5.5) | 46 (6.4) | 34 (8.1) | 42 (2.8) | 47 (3.2) | 52 (4.8) | 32 (8.4) | 67 (3.4) | 77 (3.8) | 98 (5.4) | 66 (8.3) |
| 45-49 | 2 (0.5) | 1 (0.2) | 3 (0.4) | 2 (0.5) | 6 (0.4) | 10 (0.7) | 10 (0.9) | 3 (0.8) | 8 (0.4) | 11 (0.5) | 13 (0.7) | 5 (0.6) |
| 50+ | 0 (0.0) | 1 (0.2) | 1 (0.1) | 1 (0.2) | 1 (0.1) | 0 (0.0) | 1 (0.1) | 0 (0.0) | 1 (0.1) | 1 (0.0) | 2 (0.1) | 1 (0.1) |
| missing | 1 (0.2) | 2 (0.4) | 3 (0.4) | 1 (0.2) | 3 (0.2) | 1 (0.1) | 0 (0.0) | 0 (0.0) | 4 (0.2) | 3 (0.1) | 3 (0.2) | 1 (0.1) |
| **Marital status** N (%) |  |  |  |  |  |  |  |  |  |  |  |  |
| Married/ Co-habiting | 434 (98.9) | 532 (98.5) | 701 (97.5) | 408 (97.6) | 1403 (92.2) | 1360 (92.1) | 1001 (92.8) | 360 (94.7) | 1837 (93.6) | 1892 (93.8) | 1702 (94.7) | 768 (96.2) |
| Never married (or single) | 5 (1.1) | 8 (1.5) | 15 (2.1) | 8 (1.9) | 94 (6.2) | 94 (6.4) | 64 (5.9) | 16 (4.2) | 99 (5.0) | 102 (5.1) | 79 (4.4) | 24 (3.0) |
| Separated/Divorced/Widowed | 0 (0.0) | 0 (0.0) | 3 (0.4) | 2 (0.5) | 24 (1.6) | 21 (1.4) | 14 (1.3) | 4 (1.1) | 24 (1.2) | 21 (1.0) | 17 (0.9) | 6 (0.8) |
| missing | 0 (0.0) | 0 (0.0) | 0 (0.0) | 0 (0.0) | 2 (0.1) | 1 (0.1) | 0 (0.0) | 0 (0.0) | 2 (0.1) | 1 (0.0) | 0 (0.0) | 0 (0.0) |
| **Education** N (%) |  |  |  |  |  |  |  |  |  |  |  |  |
| Higher | 21 (4.8) | 22 (4.1) | 28 (3.9) | 15 (3.6) | 209 (13.7) | 207 (14.0) | 133 (12.3) | 38 (10.0) | 230 (11.7) | 229 (11.4) | 161 (9.0) | 53 (6.6) |
| None | 272 (62.0) | 348 (64.4) | 466 (64.8) | 269 (64.4) | 118 (7.8) | 110 (7.5) | 76 (7.0) | 25 (6.6) | 390 (19.9) | 458 (22.7) | 542 (30.1) | 294 (36.8) |
| Primary | 68 (15.5) | 84 (15.6) | 116 (16.1) | 76 (18.2) | 781 (51.3) | 742 (50.3) | 583 (54.0) | 216 (56.8) | 849 (43.3) | 826 (41.0) | 699 (38.8) | 292 (36.6) |
| Secondary | 78 (17.8) | 86 (15.9) | 109 (15.2) | 58 (13.9) | 413 (27.1) | 416 (28.2) | 287 (26.6) | 101 (26.6) | 491 (25.0) | 502 (24.9) | 396 (22.0) | 159 (19.9) |
| Missing | 0 (0.0) | 0 (0.0) | 0 (0.0) | 0 (0.0) | 2 (0.1) | 1 (0.1) | 0 (0.0) | 0 (0.0) | 2 (0.1) | 1 (<0.1) | 0 (0.0) | 0 (0.0) |
| **Occupation** N (%) |  |  |  |  |  |  |  |  |  |  |  |  |
| Business | 0 (0.0) | 0 (0.0) | 0 (0.0) | 0 (0.0) | 141 (9.3) | 140 (9.5) | 94 (8.7) | 41 (10.8) | 141 (7.2) | 140 (6.9) | 94 (5.2) | 41 (5.1) |
| Construction | 0 (0.0) | 0 (0.0) | 0 (0.0) | 0 (0.0) | 0 (0.0) | 0 (0.0) | 0 (0.0) | 0 (0.0) | 0 (0.0) | 0 (0.0) | 0 (0.0) | 0 (0.0) |
| Factory | 0 (0.0) | 0 (0.0) | 0 (0.0) | 0 (0.0) | 29 (1.9) | 28 (1.9) | 30 (2.8) | 15 (3.9) | 29 (1.5) | 28 (1.4) | 30 (1.7) | 15 (1.9) |
| Housewife | 384 (87.5) | 478 (88.5) | 643 (89.4) | 373 (89.2) | 803 (52.7) | 772 (52.3) | 567 (52.5) | 204 (53.7) | 1187 (60.5) | 1250 (62.0) | 1210 (67.3) | 577 (72.3) |
| Informal - Employment | 0 (0.0) | 0 (0.0) | 0 (0.0) | 0 (0.0) | 200 (13.1) | 206 (14.0) | 144 (13.3) | 47 (12.4) | 200 (10.2) | 206 (10.2) | 144 (8.0) | 47 (5.9) |
| Large-scale agriculture | 4 (0.9) | 4 (0.7) | 5 (0.7) | 4 (1.0) | 0 (0.0) | 0 (0.0) | 0 (0.0) | 0 (0.0) | 4 (0.2) | 4 (0.2) | 5 (0.3) | 4 (0.5) |
| Market trader | 17 (3.9) | 17 (3.1) | 22 (3.1) | 12 (2.9) | 177 (11.6) | 164 (11.1) | 124 (11.5) | 39 (10.3) | 194 (9.9) | 181 (9.0) | 146 (8.1) | 51 (6.4) |
| Other (specify) | 20 (4.6) | 25 (4.6) | 33 (4.6) | 25 (6.0) | 18 (1.2) | 15 (1.0) | 10 (0.9) | 3 (0.8) | 38 (1.9) | 40 (2.0) | 43 (2.4) | 28 (3.5) |
| Professional | 8 (1.8) | 10 (1.9) | 10 (1.4) | 3 (0.7) | 125 (8.2) | 118 (8.0) | 86 (8.0) | 27 (7.1) | 133 (6.8) | 128 (6.3) | 96 (5.3) | 30 (3.8) |
| Student | 6 (1.4) | 6 (1.1) | 6 (0.8) | 1 (0.2) | 28 (1.8) | 31 (2.1) | 23 (2.1) | 4 (1.1) | 34 (1.7) | 37 (1.8) | 29 (1.6) | 5 (0.6) |
| missing | 0 (0.0) | 0 (0.0) | 0 (0.0) | 0 (0.0) | 2 (0.1) | 2 (0.1) | 1 (0.1) | 0 (0.0) | 2 (0.1) | 2 (0.1) | 1 (0.1) | 0 (0.0) |
| **Mother started employment / returned to school since giving birth** N (%) |  |  |  |  |  |  |  |  |  |  |  |  |
| Yes | 110 (25.1) | 132 (24.4) | 302 (42.0) | 177 (42.3) | 468 (30.7) | 730 (49.5) | 631 (58.4) | 230 (60.5) | 578 (29.5) | 863 (42.8) | 933 (51.9) | 407 (51.0) |
| **Household composition** N (%) |  |  |  |  |  |  |  |  |  |  |  |  |
| Total Number of people in the household | 13.0 (9.0, 18.0) | 13.0 (9.0, 19.0) | 13.0 (10.0, 19.0) | 13.0 (9.0, 18.0) | 4.0 (3.0, 6.0) | 4.0 (3.0, 6.0) | 4.0 (3.0, 7.0) | 5.0 (3.0, 6.0) | 5.0 (3.0, 9.0) | 5.0 (3.0, 9.0) | 7.0 (4.0, 12.0) | 8.0 (5.0, 13.0) |
| Total number of people over 18 | 6 (4, 10) | 6 (5, 10) | 6 (4, 9) | 6 (4, 10) | 2 (2, 3) | 2 (2, 3) | 2 (2, 3) | 2 (2, 3) | 2.0 (2.0, 5.0) | 3.0 (2.0, 5.0) | 3.0 (2.0, 6.0) | 4.0 (2.0, 6.0) |
| Total number of people under 18 | 6.0 (4.0, 10.0) | 6.0 (4.0, 10.0) | 6.0 (4.0, 10.0) | 6.0 (4.0, 10.0) | 2.0 (1.0, 3.0) | 2.0 (1.0, 3.0) | 2.0 (1.0, 3.0) | 2.0 (1.0, 3.0) | 2.0 (1.0, 4.0) | 3.0 (1.0, 5.0) | 3.0 (2.0, 6.0) | 4.0 (2.0, 7.0) |
| Father living with the child | 341 (77.7) | 433 (80.2) | 552 (76.8) | 330 (78.9) | 1279 (84.0) | 1245 (84.3) | 852 (78.9) | 305 (80.3) | 1620 (82.6) | 1678 (83.2) | 1404 (78.0) | 635 (79.6) |
| Missing | 12 (2.7) | 13 (2.4) | 23 (3.2) | 8 (1.9) | 24 (1.6) | 23 (1.6) | 43 (4.0) | 12 (3.2) | 36 (1.8) | 36 (1.8) | 66 (3.7) | 20 (2.5) |
| Number of mother who were pregnant at the visit | 6 (1.4) | 15 (2.8) | 149 (20.6) | 55 (13.2) | 8 (0.5) | 38 (2.6) | 83 (7.7) | 25 (6.6) | 14 (0.7) | 53 (2.6) | 231 (12.8) | 80 (10.0) |
| **Maternal BMI, median (IQR)** | 21.5 [19.2-25.3] | 21.3 [18.7-25.0] | 22.0 [19.2-25.2] | 21.8 [19.4-24.5] | 23.6 [20.9-27.5] | 23.5 [20.3-27.9] | 23.6 [20.4-28.2] | 24.6 [21.0-28.9] | 23.3 [20.4-27.0] | 22.9 [19.8-26.9] | 22.9 [19.8-27.1] | 22.9 [19.9-26.7] |
| **Maternal BMI Category** N (%) |  |  |  |  |  |  |  |  |  |  |  |  |
| <18.5 | 80 (18.2) | 118 (21.9) | 121 (16.8) | 75/418 (17.9) | 111/1515 (7.3) | 161/1458 (11.0) | 113/1053 (10.7) | 33/363 (9.1) | 191/1954 (9.8) | 279 (14.0) | 234 (13.2) | 108/781 (13.8) |
| 18.5-24.9 | 237 (54.0) | 287 (53.1) | 400 (55.7) | 252/418 (60.3) | 814/1515 (53.7) | 720/1458 (49.4) | 493/1053 (46.8) | 157/363 (43.3) | 1051/1954 (53.8) | 1007 (50.4) | 893 (50.4) | 409/781 (52.4) |
| 25-29.9 | 77 (17.5) | 88 (16.3) | 131 (18.2) | 63/418 (15.1) | 367/1515 (24.2) | 327/1458 (22.4) | 255/1053 (24.2) | 97/363 (26.7) | 444/1954 (22.7) | 415 (20.8) | 386 (21.8) | 160/781 (20.5) |
| 30 | 42 (9.6) | 44 (8.1) | 55 (7.6) | 23/418 (5.5) | 216/1515 (14.3) | 244/1458 (16.7) | 188/1053 (17.9) | 71/363 (19.6) | 258/1954 (13.2) | 288 (14.4) | 243 (13.7) | 94/781 (12.0) |
| Missing | 3 (0.7) | 3 (0.6) | 12 (1.7) | 5/418 (1.2) | 7/1515 (0.5) | 6/1458 (0.4) | 4/1053 (0.4) | 5/363 (1.4) | 10/1954 (0.5) | 9 (0.5) | 16 (0.9) | 10/781 (1.3) |
| **MUAC- median (IQR)** | 27.3 [24.8-30.3] | 27.0 [24.4-30.3] | 27.0 [24.6-30.0] | 27.1 [24.7-30.0] | 27.1 [24.6-30.2] | 27.1 [24.5-30.5] | 27.2 [24.5-31.0] | 27.6 [24.6-31.5] | 27.1 [24.6-30.2] | 27.1 [24.5-30.5] | 27.1 [24.5-30.4] | 27.3 [24.7-30.5] |
| **MUAC Category** N (%) |  |  |  |  |  |  |  |  |  |  |  |  |
| Underweight (<23.0cm) | 47 (10.7) | 61 (11.3) | 83 (11.5) | 33/418 (7.9) | 157/1515 (10.4) | 191/1458 (13.1) | 127/1053 (12.1) | 39/363 (10.7) | 204/1954 (10.4) | 252 (12.6) | 210 (11.8) | 72/781 (9.2) |
| Normal weight (23.0-31.9cm) | 313 (71.3) | 394 (73.0) | 525 (73.0) | 326/418 (78.0) | 1120/1515 (73.9) | 1003/1458 (68.8) | 721/1053 (68.5) | 240/363 (66.1) | 1433/1954 (73.3) | 1397 (69.9) | 1246 (70.3) | 566/781 (72.4) |
| Overweight/obese (≥32.0cm) | 78 (17.8) | 85 (15.7) | 107 (14.9) | 56/418 (13.4) | 237/1515 (15.6) | 262/1458 (18.0) | 202/1053 (19.2) | 81/363 (22.3) | 315/1954 (16.1) | 347 (17.4) | 309 (17.4) | 137/781 (17.5) |
| Missing | 1 (0.2) | 0 (0.0) | 4 (0.6) | 3/418 (0.7) | 1/1515 (0.1) | 2/1458 (0.1) | 3/1053 (0.3) | 3/363 (0.8) | 2/1954 (0.1) | 2 (0.1) | 7 (0.4) | 6/781 (0.8) |
| BP Category N (%) |  |  |  |  |  |  |  |  |  |  |  |  |
| Normal | 328 (74.7) | 438 (81.1) | 637 (88.6) | 351 (84.0) | 1057 (69.8) | 1109 (76.1) | 811 (77.0) | 271 (74.7) | 1385 (70.9) | 1547 (77.4) | 1448 (81.7) | 622 (79.6) |
| Elevated | 18 (4.1) | 25 (4.6) | 22 (3.1) | 12 (2.9) | 90 (5.9) | 73 (5.0) | 40 (3.8) | 19 (5.2) | 108 (5.5) | 98 (4.9) | 62 (3.5) | 31 (4.0) |
| Stage 1 hypertension | 69 (15.7) | 63 (11.7) | 47 (6.5) | 41 (9.8) | 288 (19.0) | 224 (15.4) | 166 (15.8) | 57 (15.7) | 357 (18.3) | 287 (14.4) | 213 (12.0) | 98 (12.5) |
| Stage 2 hypertension | 24 (5.5) | 13 (2.4) | 10 (1.4) | 14 (3.3) | 79 (5.2) | 51 (3.5) | 34 (3.2) | 14 (3.9) | 103 (5.3) | 64 (3.2) | 44 (2.5) | 28 (3.6) |
| missing | 0 (0.0) | 1 (0.2) | 3 (0.4) | 0 (0.0) | 1 (0.1) | 1 (0.1) | 2 (0.2) | 2 (0.6) | 1 (0.1) | 2 (0.1) | 5 (0.3) | 2 (0.3) |
| **Cardiology assessment** |  |  |  |  |  |  |  |  |  |  |  |  |
| Pulse wave velocity | - | 7.1 (6.5, 8.2) |  | - | - | 7.1 (6.6, 7.9) |  | - | - | 7.1 (6.5, 7.9) |  | - |
| Cardiac Output | - | 4.2 (3.4, 4.9) |  | - | - | 4.8 (4.0, 5.8) |  | - | - | 4.7 (3.8, 5.6) |  | - |
| Systemic Vascular Resistance | - | 1578.9 (1355.8, 2020.9) |  | - | - | 1401.5 (1150.6, 1774.8) |  | - | - | 1456.2 (1201.4, 1840.3) |  | - |
| **Nutrition Status** N (%) |  |  |  |  |  |  |  |  |  |  |  |  |
| Meets minimum dietary diversity | 325 (74.0) | 439 (81.3) | 610 (84.8) | 360/418 (86.1) | 792/1515 (52.3) | 825/1458 (56.6) | 607/1053 (57.6) | 207/363 (57.0) | 1117/1954 (57.2) | 1264/1998 | 1217/1772 () | 567/781 (72.6) |
| missing | 0 | 0 | 0 | 0 | 0 | 0 | 0 | 0 | 0 | 0 | 0 | 0 |
| **Mental health** N (%) |  |  |  |  |  |  |  |  |  |  |  |  |
| Number of participants who screened positive for WHODAS | 0/439 (0.0) | 0/3 (0) | 1/370 (0.3) | 1/288 (0.3) | 21/ 1515 (1.4) | 46/704 (6.5) | 50/847 (5.9) | 20/323 (6.2) | 21/1954 (1.1) | 46/707 (6.5) | 51/1217 (4.2) | 21/611 (3.4) |
| Number of participants who screened positive for anxiety | 5/438 (1.1) | 1/370 (0.3) | 2/365 (0.5) | 0/287 (0) | 13/544 (2.4) | 46/866 (5.3) | 29/771 (3.8) | 5/251 (2.0) | 18/982 (1.8) | 47/1236 (3.8) | 31/1136 (2.7) | 5/538 (0.9) |
| Number of participants who screened positive for depression | 3/438 (0.7) | 0/370 (0) | 3/365 (0.8) | 1/287 (0.3) | 7/544 (1.3) | 21/866 (2.4) | 12/771 (1.6) | 4/251 (1.6) | 10/982 (1.0) | 21/1236 (1.7) | 15/1136 (1.3) | 5/538 (0.9) |
| Number of participants who screened positive for post traumatic stress | 0/3 (0) | 2/365 (0.5) | 1/22 (4.5) | 1/16 (6.3) | 2/ 29 (6.9) | 24/473 (5.1) | 13/121 (10.7) | 3/31 (9.7) | 2/32 (6.3) | 26/838 (3.1) | 14/143 (9.8) | 4/47 (8.5) |
| Number of participants who had suicidal thoughts | 5/438 (1.1) | 2/370 (0.5) | 4/365 (1.1) | 2/287 (0.7) | 14/544 (2.6) | 60/866 (6.9) | 46/771 (6.0) | 16/251 (6.4) | 19/982 (1.9) | 62/1236 (5.0) | 50/1136 (4.4) | 18/538 (3.3) |

Table S3 Children clinical profile by visit

|  | **The Gambia** | | | | **Kenya** | | | | **All countries** | | | |
| --- | --- | --- | --- | --- | --- | --- | --- | --- | --- | --- | --- | --- |
|  | **Visit 1 (6 wk-6 months after birth)** | **Visit 2 (12 months after birth)** | **Visit 3 (24 months after birth)** | **Visit 4 (36 months after birth)** | **Visit 1 (6 wk-6 months after birth)** | **Visit 2 (12 months after birth)** | **Visit 3 (24 months after birth)** | **Visit 4 (36 months after birth)** | **Visit 1 (6 wk-6 months after birth)** | **Visit 2 (12 months after birth)** | **Visit 3 (24 months after birth)** | **Visit 4 (36 months after birth)** |
| **Number of children** | 433 | 529 | 692 | 397 | 1511 | 1453 | 1076 | 391 | 1944 | 1982 | 1768 | 788 |
| Girls N (%) | 212 (49.0) | 264 (49.9) | 348 (50.3) | 197 (49.6) | 751 (49.7) | 710 (48.9) | 522 (48.5) | 170 (43.5) | 963 (49.5) | 974 (49.1) | 870 (49.2) | 367 (46.6) |
| Boys N (%) | 219 (50.6) | 263 (49.7) | 344 (49.7) | 200 (50.4) | 757 (50.1) | 737 (50.7) | 548 (50.9) | 218 (55.8) | 976 (50.2) | 1000 (50.5) | 892 (50.5) | 418 (53.0) |
| Missing | 2 (0.5) | 2 (0.4) | 0 (0.0) | 0 (0.0) | 3 (0.2) | 6 (0.4) | 6 (0.6) | 3 (0.8) | 5 (0.3) | 8 (0.4) | 6 (0.3) | 3 (0.6) |
| **GA at birth (weeks [IQR])** | 39.1 (37.4, 40.6) | 39.0 (37.3, 40.6) | 39.1 (37.7, 40.6) | 39.0 (37.5, 40.1) | 39.1 (37.3, 40.6) | 39.1 (37.6, 40.6) | 39.1 (37.3, 40.6) | 39.1 (37.9, 40.6) | 39.1 (37.3, 40.6) | 39.1 (37.4, 40.6) | 39.1 (37.4, 40.6) | 39.1 (37.6, 40.3) |
| **Age(months) - Median (IQR)** | 5.5 [5.0-5.8] | 11.8 [11.1-12.6] | 24.0 [23.5-24.6] | 35.9 [35.4-36.4] | 3.3 [3.0-4.0] | 11.1 [11.0-11.7] | 23.1 [23.0-23.6] | 35.1 [35.0-35.5] | 3.5 [3.1-5.0] | 11.2 [11.0-12.0] | 23.4 [23.0-24.2] | 35.4 [35.1-36.1] |
| **Child health** N (%) |  |  |  |  |  |  |  |  |  |  |  |  |
| Hospital admission |  |  |  |  |  |  |  |  |  |  |  |  |
| yes | 9/433 (2.1) | 10/529 (1.9) | 23/692 (3.3) | 8/397 (2.0) | 37/1503 (2.5) | 95/1435 (6.6) | 94/1051 (8.9) | 26/374 (7.0) | 46/1936 (2.4) | 105/1964 (5.3) | 117/1743 (6.7) | 34/771 (4.4) |
| missing | 3/433 (0.7) | 6/529 (1.1) | 8/692 (1.2) | 7/397 (1.8) | 6/1503 (0.4) | 7/1435 (0.5) | 5/1051 (0.5) | 0/374 (0.0) | 9/1936 (0.5) | 13/1964 (0.7) | 13/1743 (0.7) | 16/771 (2.1) |
| Hospital stay length- days- median (IQR) | 4.5 [2.3-5.0] | 5.0 [3.3-8.0] | 3.0 [2.0-7.5] | 8.5 [6.0-14.8] | 9.0 [4.0-14.0] | 5.0 [3.0-7.0] | 4.0 [3.0-7.0] | 3.0 [3.0-6.8] | 6.0 [3.0-7.3] | 5.0 [3.0-7.0] | 4.0 [2.0-7.0] | 4.5 [3.0-8.8] |
| Malaria test | 14/433 (3.2) | 32/529 (6.0) | 77/692 (11.1) | 38/397 (9.6) | 192/1503 (12.8) | 729/1435 (50.8) | 709/1051 (67.5) | 241/374 (64.4) | 206/1936 (10.6) | 761/1964 (38.7) | 786/1743 (45.1) | 279/771 (36.2) |
| Test result positive | 0/433 (0.0) | 0/529 (0.0) | 0/692 (0.0) | 2/397 (0.5) | 12/1503 (0.8) | 76/1435 (5.3) | 102/1051 (9.7) | 49/374 (13.1) | 12/1936 (0.6) | 76/1964 (3.9) | 102/1743 (5.9) | 51/771 (6.6) |
| Child has difficulty seeing | - | 5/529 (0.9) | 0/692 (0.0) | 1/397 (0.3) | - | 7/1435 (0.5) | 4/1051 (0.4) | 2/374 (0.5) | - | 12/1964 (0.6) | 4/1743 (0.2) | 3/771 (0.4) |
| Child has difficulty hearing | - | 4/529 (0.8) | 0/692 (0.0) | 0/397 (0.0) | - | 12/1435 (0.8) | 3/1051 (0.3) | 0/374 (0.0) | - | 16/1964 (0.8) | 3/1743 (0.2) | 0/771 (0.0) |
| Child cough when no fever or illness | - | 94/529 (17.8) | 86/692 (12.4) | 31/397 (7.8) | - | 353/1435 (24.6) | 223/1051 (21.2) | 93/374 (24.9) | - | 447/1964 (22.8) | 309/1743 (17.7) | 124/771 (16.1) |
| Child has wheezing or whistling in the chest | - | 22/529 (4.2) | 26/692 (3.8) | 9/397 (2.3) | - | 134/1435 (9.3) | 143/1051 (13.6) | 45/374 (12.0) | - | 156/1964 (7.9) | 169/1743 (9.7) | 54/771 (7.0) |
| **Blood Pressure** N (%) |  |  |  |  |  |  |  |  |  |  |  |  |
| BP ≥90th percentile | - | 183/529 (34.6) | 515/692 (74.4) | 223/397 (56.2) | - | 194/1435 (13.5) | 607/1051 (57.8) | 246/374 (65.8) | - | 377/1964 (19.2) | 1122/1743 (64.4) | 469/771 (60.8) |
| BP <10th percentile | - | 18/529 (3.4) | 0/692 (0.0) | 1/397 (0.3) | - | 228/1435 (15.9) | 7/1051 (0.7) | 1/374 (0.3) | - | 246/1964 (12.5) | 7/1743 (0.4) | 2/771 (0.3) |
| **Child nutrition Status** N (%) |  |  |  |  |  |  |  |  |  |  |  |  |
| children breastfed exclusively | 244/433 (56.4) | - | - | - | 1187/1511 (78.6) | - | - | - | 1431/1944 (73.6) | - | - | - |
| never breastfed | 4/433 (0.9) | - | - | - | 9/1511 (0.6) | - | - | - | 13/1944 (0.7) | - | - | - |
| Still breastfeeding (0) | - | 69/529 (13.0) | 23/692 (3.3) | 2/397 (0.5) | - | 34/1435 (2.4) | 8/1051 (0.8) | 0/374 (0.0) | - | 103/1964 (5.2) | 31/1743 (1.8) | 2/771 (0.3) |
| stopped exclusive breastfeeding 0-3 month (1) | - | 48/529 (9.1) | 55/692 (7.9) | 36/397 (9.1) | - | 68/1435 (4.7) | 52/1051 (4.9) | 16/374 (4.3) | - | 116/1964 (5.9) | 107/1743 (6.1) | 52/771 (6.7) |
| stopped exclusive breastfeeding 4-6 month (2) | - | 282/529 (53.3) | 392/692 (56.6) | 215/397 (54.2) | - | 1044/1435 (72.8) | 732/1051 (69.6) | 259/374 (69.3) | - | 1326/1964 (67.5) | 1124/1743 (64.5) | 474/771 (61.5) |
| stopped exclusive breastfeeding at 6 month or later | - | 129/529 (24.4) | 215/692 (31.1) | 142/397 (35.8) | - | 289/1435 (20.1) | 256/1051 (24.4) | 98/374 (26.2) | - | 418/1964 (21.3) | 471/1743 (27.0) | 240/771 (31.1) |
| missing | 0/433 (0.0) | 1/529 (0.2) | 3/692 (0.4) | 2/397 (0.5) | 1/1511 (0.1) | 0/1435 (0.0) | 3/1051 (0.3) | 1/374 (0.3) | 1/1944 (0.1) | 1/1964 (0.1) | 10/1743 (0.6) | 3/771 (0.4) |
| Height/length for age (stunting) (z-score) | -0.17 (-1.10,0.49) | -0.99 (-1.77,-0.16) | -1.46 (-2.15,-0.83) | -1.21 (-1.73,-0.56) | -0.46 (-1.32,0.39) | -0.98 (-1.79,-0.16) | -1.20 (-2.13,-0.32) | -0.86 (-1.58,-0.13) | -0.40 (-1.25,0.43) | -0.99 (-1.77,-0.16) | -1.34 (-2.13,-0.53) | -1.11 (-1.67,-0.33) |
| Stunted N (%) | 34/433 (7.9) | 86/529 (16.3) | 193/692 (27.9) | 63/397 (15.9) | 168/1503 (11.2) | 259/1435 (18.1) | 284/1051 (27.0) | 59/374 (15.8) | 202/1936 (10.4) | 345/1964 (17.6) | 477/1743 (27.4) | 122/771 |
| Missing | 5/433 (1.2) | 7/529 (1.3) | 18/692 (2.6) | 8/397 (2.0) | 22/1511 (1.5) | 34/1453 (2.3) | 39/1076 (3.6) | 24/391 (6.1) | 27/1944 (1.4) | 41/1982 (2.1) | 57/1768 (3.2) | 32/788 (4.1) |
| Weight for height/length (wasting) (z-score) | -0.77 (-1.58,0.01) | -1.0 (-1.81,-0.21) | -0.72 (-1.42,0.02) | -0.80 (-1.38,-0.23) | 0.10 (-0.72,0.89) | -0.21 (-1.03,0.58) | -0.42 (-1.17,0.33) | -0.65 (-1.35,0.10) | -0.10 (-0.94,0.75) | -0.43 (-1.27,0.37) | -0.54 (-1.30,0.22) | -0.73 (-1.35,-0.09) |
| Wasted N (%) | 68/433 (15.7) | 108/529 (20.4) | 76/692 (11.0) | 44/397 (11.1) | 74/1503 (4.9) | 102/1435 (7.1) | 87/1051 (8.3) | 30/374 (8.0) | 142/1936 (7.3) | 210/1964 (10.7) | 163/1743 (9.4) | 74/771 (9.6) |
| Missing | 7/433 (1.6) | 7/529 (1.3) | 18/692 (2.6) | 7/397 (1.8) | 23/1511 (1.5) | 34/1453 (2.3) | 39/1076 (3.6) | 24/391 (6.1) | 30/1944 (1.5) | 41/1982 (2.1) | 57/1768 (3.2) | 31/788 (3.9) |
| **MUAC z score** | -0.38 (-1.04,0.25) | -0.68 (-1.30,0.03) | -0.98 (-1.58,-0.36) | -0.84 (-1.38,-0.29) | 0.24 (-0.51,0.98) | -0.10 (-0.83,0.72) | -0.40 (-1.05,0.28) | -0.47 (-1.11,0.34) | 0.05 (-0.64,0.83) | -0.24 (-1.0,0.56) | -0.61 (-1.31,0.08) | -0.64 (-1.26,-0.04) |
| MUAC under threshold | 27/433 (6.2) | 49/529 (9.3) | 93/692 (13.4) | 44/397 (11.1) | 47/1503 (3.1) | 73/1435 (5.1) | 52/1051 (5.0) | 24/374 (6.4) | 74/1936 (3.8) | 122/1964 (6.2) | 145/1743 (8.3) | 68/771 (8.8) |
| <11.5cm (severe malnutrition) | 23/433 (5.3) | 16/529 (3.0) | 10/692 (1.4) | 2/397 (0.5) | 62/1503 (4.1) | 19/1435 (1.3) | 9/1051 (0.9) | 3/374 (0.8) | 85/1936 (4.4) | 35/1964 (1.8) | 19/1743 (1.1) | 5/771 (0.6) |
| Between 11.5 and 12.5cm (moderate malnutrition) | 62/433 (14.3) | 60/529 (11.3) | 47/692 (6.8) | 7/397 (1.8) | 176/1503 (11.7) | 80/1435 (5.6) | 22/1051 (2.1) | 2/374 (0.5) | 238/1936 (12.3) | 140/1964 (7.1) | 69/1743 (4.0) | 9/771 (1.2) |
| Missing | 0/433 (0.0) | 2/529 (0.4) | 6/692 (0.9) | 7/397 (1.8) | 10/1511 (0.7) | 21/1453 (1.4) | 29/1076 (2.7) | 19/391 (4.9) | 10/1944 (0.5) | 23/1982 (1.2) | 35/1768 (2.0) | 26/788 (3.3) |
| Neuro Assessment N (%) |  |  |  |  |  |  |  |  |  |  |  |  |
| number of children assessed with MDAT | 427 | 523 | 678 | 390 | 1493 | 1418 | 1015 | 369 | 1920 | 1941 | 1693 | 759 |
| Screened positive MDAT <-1SD | 88 (20.6) | 48 (9.2) | 85 (12.5) | 55 (14.1) | 147 (9.8) | 128 (9.0) | 105 (10.3) | 29 (7.9) | 235 (12.2) | 176 (9.1) | 190 (11.2) | 84 (11.1) |
| Screened positive MDAT <-2SD | 16 (3.7) | 11 (2.1) | 12 (1.8) | 8 (2.1) | 30 (2.0) | 41 (2.9) | 26 (2.6) | 16 (4.3) | 46 (2.4) | 52 (2.7) | 38 (2.2) | 24 (3.2) |
| number of children assessed with OMCI | 428 | - | 678 | - | 1498 | - | 1018 | - | 1917 | - | 1696 | - |
| number of children assessed with GMA video | 433 | - | - | - | 1501 | - | - | - | 1934 | - | - | - |
| number of children assessed with Family Care Indicators questionnaire | - | 528 | 690 | 395 | - | 1435 | 1051 | 374 | - | 1963 | 1741 | 769 |
| number of children assessed with NDST | - | - | 689 | 394 | - | - | 1050 | 374 | - | - | 1739 | 768 |
| At risk of developmental delay | - | - | 15/689 (2.2) | 3/395 (0.8) | - | - | 57/1051 (5.4) | 25/374 (6.7) | - | - | 72/1740 (4.1) | 28/769 (3.6) |
| Screened positive NDST |  |  |  |  |  |  |  |  |  |  |  |  |
| number of children assessed with epilepsy questionnaire | - | - | 689 | 394 | - | - | 1051 | 374 | - | - | 1739 | 768 |
| Screened positive epilepsy | - | - | 3 (0.4) | 1 (0.3) | - | - | 35/1051 (3.3) | 16/374 (4.3) | - | - | 38/1739 (2.2) | 17/768 (2.2) |
| Number of children flagged during the study******* |  |  | 16/689 (2.3) | 4/394 (1.0) |  |  | 59/1050 (5.6) | 25/369 (6.8) |  |  | 75/1739 (4.3) | 29/768 (3.8) |
| number of children assessed with PedSQL | - | - | 16/16 | 4/4 | - | - | 59/59 | 25/25 | - | - | 75/75 | 29/29 |
| number of children Screened positive for MCHAT | - | - | 9/16 (56.3) | 1/4 (25.0) | - | - | 14/59 (23.7) | 3/25 (12.0) | - | - | 23/75 (30.7) | 4/29 (13.8) |
| number of children Screened positive for CARDIF |  | - | 15/16 (93.8) | 2/4 (50.0) | - | - | 52/59 (88.1) | 23/25 (92.0) | - | - | 67/75 (89.3) | 25/29 (86.2) |
| **Number of phone interview** | - | - | - | - | 8/1511 (0.5) | 18/1453 (1.2) | 26/1076 (2.4) | 17/391 (4.3) | 8/1944 (0.4) | 18/1982 (0.9) | 26/1768 (1.5) | 17/788 (2.2) |
| number of children assessed with Developmental Milestones Checklist (DMC-III) | - | 0 (0.0) | - | - | 8/8 (100.0) | 18/18 (100.0) | 26/26 (100.0) | 17/17 (100.0) | 8/8 (100.0) | 18/18 (100.0) | 26/26 (100.0) | 17/17 (100.0) |
| *******Children flagged during the study for assessment with MDAT and NDST were asked the following questionnaire: PedSQL, MCHAT, and CARDIF | | | | | | | | | | | | |

**Table S4** **Pregnancy outcomes of participants recruited to quality of care and health economics sub-studies**

| **Quality of care** | The Gambia | Kenya | All countries |
| --- | --- | --- | --- |
| **Number of participants** | 437 | 695 | 1132 |
| **Controls: Uncomplicated pregnancy** N (%) | 210 (48.0) | 328 (47.2) | 538 (47.5) |
| **Cases** N (%) |  |  |  |
| Stage 2 Hypertension | 16 (3.7) | 47 (6.8) | 63 (5.6) |
| Caesarean section | 21 (4.8) | 117 (16.8) | 138 (12.2) |
| Stillbirth | 19 (4.3) | 12 (1.7) | 31 (2.7) |
| Neonatal death | 1 (0.2) | 4 (0.6) | 5 (0.4) |
| Small vulnerable newborn | 194 (44.4) | 279 (40.1) | 473 (41.8) |
| Small for gestational age <3rd percentile | 39 (8.9) | 57 (8.2) | 96 (8.5) |
| Preterm birth <33 weeks | 20 (4.6) | 33 (4.7) | 53 (4.7) |
| **Health Economics** | The Gambia | Kenya | All countries |
| **Number of participants** | 110 | 100 | 210 |
| **Controls: Uncomplicated pregnancy** N (%) | 56 (50.9) | 46 (46.0) | 102 (48.6) |
| **Cases** N (%) |  |  |  |
| Stage 2 hypertension | 2 (1.8) | 3 (3.0) | 5 (2.4) |
| Caesarean section | 5 (4.5) | 12 (12.0) | 17 (8.1) |
| Stillbirth | 7 (6.4) | 0 (0.0) | 7 (3.3) |
| Neonatal death | 0 (0.0) | 3 (3.0) | 3 (1.4) |
| Small vulnerable newborn | 35 (31.8) | 39 (39.0) | 74 (35.2) |
| Small for gestational age <3rd percentile | 13 (11.8) | 5 (5.0) | 18 (8.6) |
| Preterm birth <33 weeks | 3 (2.7) | 4 (4.0) | 7 (3.3) |

Table S5 Number of aliquots collected for each sample type by study visit

|  | Gambia | | | | Kenya | | | | Total | | | |
| --- | --- | --- | --- | --- | --- | --- | --- | --- | --- | --- | --- | --- |
| **Women** | **Visit 1 (6 wk-6 months after birth)** | **Visit 2 (12 months after birth)** | **Visit 3 (24 months after birth)** | **Visit 4 (36 months after birth)** | **Visit 1 (6 wk-6 months after birth)** | **Visit 2 (12 months after birth)** | **Visit 3 (24 months after birth)** | **Visit 4 (36 months after birth)** | **Visit 1 (6 wk-6 months after birth)** | **Visit 2 (12 months after birth)** | **Visit 3 (24 months after birth)** | **Visit 4 (36 months after birth)** |
| Whole blood | 426 | 520 | 660 | 383 | 985 | 1355 | 935 | 319 | 1411 | 1875 | 1595 | 702 |
| Blood spot |  | 519 |  |  |  | 1161 |  |  |  | 1680 |  |  |
| Serum | 2033 | 2347 | 2956 | 1750 | 5325 | 7321 | 5041 | 1700 | 7358 | 9668 | 7997 | 3450 |
| Plasma | 2260 | 2555 | 3239 | 1883 | 5615 | 7768 | 5422 | 1826 | 7875 | 10323 | 8661 | 3709 |
| Buffy coat | 426 | 520 | 659 | 383 | 985 | 1355 | 934 | 319 | 1411 | 1875 | 1593 | 702 |
| Urine | 772 | 1072 | 1363 | 0 | 2020 | 2792 | 9160 | 0 | 2792 | 3864 | 10523 | 0 |
| Vaginal swab - biochemistry | 1507 |  |  |  |  |  |  |  | 1507 |  |  |  |
| Vaginal swab - microbiome | 542 |  |  |  |  |  |  |  | 542 |  |  |  |
| Breastmilk (Gambia only) | 1986 |  |  |  |  |  |  |  | 1986 |  |  |  |
| **Children** | **Visit 1** | **Visit 2** | **Visit 3** | **Visit 4** | **Visit 1** | **Visit 2** | **Visit 3** | **Visit 4** | **Visit 1** | **Visit 2** | **Visit 3** | **Visit 4** |
| Blood spot | 429 | 519 | 507 | 188 | 1290 | 373 | 188 | 47 | 1719 | 892 | 695 | 235 |
| Serum |  | 2 | 263 | 323 |  | 1472 | 1184 | 469 |  | 1474 | 1447 | 792 |
| Plasma |  | 2 | 350 | 433 |  | 2169 | 1765 | 706 |  | 2171 | 2115 | 1139 |
| Buffy coat |  | 1 | 170 | 188 |  | 771 | 628 | 247 |  | 772 | 798 | 435 |
| Stool | 190 |  | 82 |  | 646 |  | 196 |  | 836 |  | 278 |  |
