## Supplementary material for "Cohort Profile: PRECISE-DYAD: a prospective cohort study linking maternal and infant health trajectories in sub-Saharan Africa": Figure S1

Figure S1a: Participants enrolment in PRECISE-DYAD study in Kenya

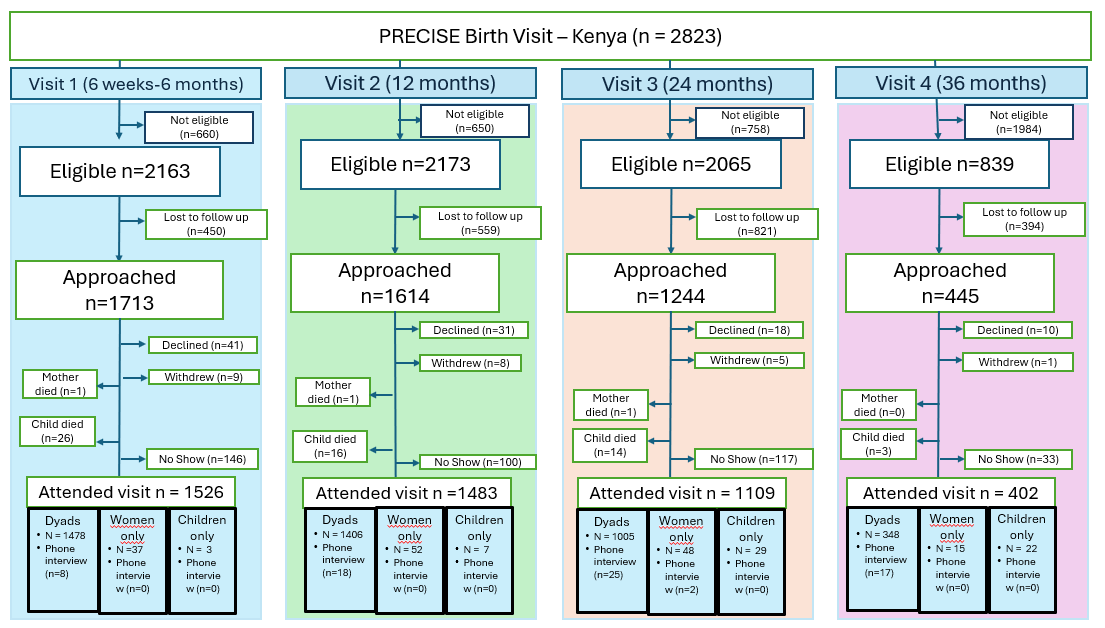

Figure S1b: Participants enrolment in PRECISE-DYAD study in The Gambia

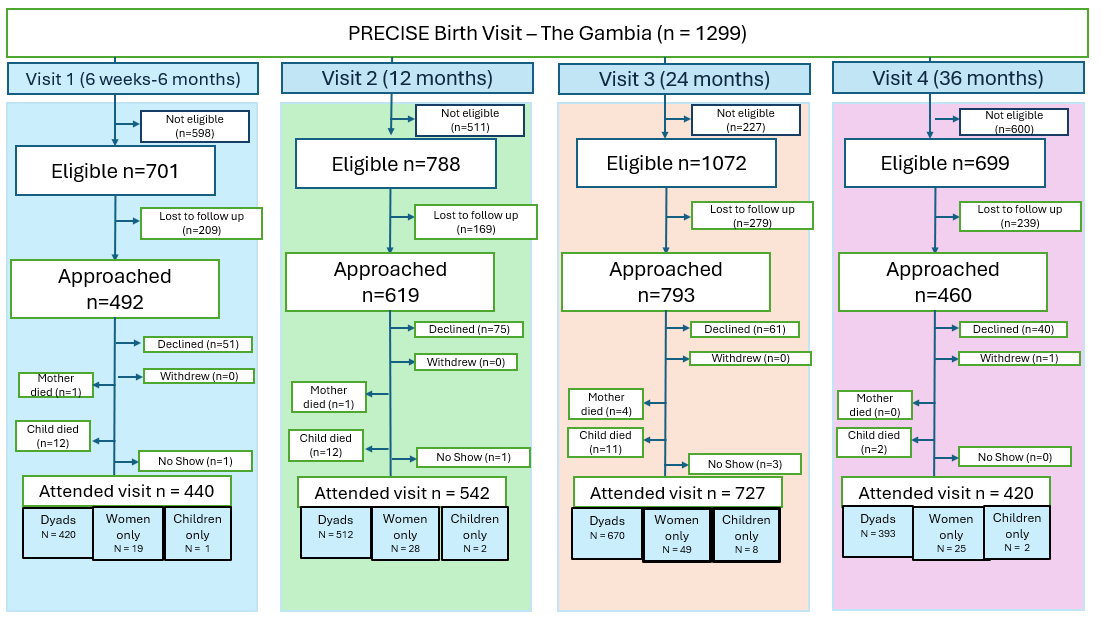

The flow chart shows the number of participants who were eligible, approached, and attended the study at each visit. Participants were enrolled at any point during the follow-up period when eligible for a PRECISE-DYAD visit (i.e., when they reached the appropriate age for that visit). Eligible participants who couldn’t be contacted (approached) were considered as lost to follow up. When the participants were approached, they had the options to decline, withdraw or take an appointment when they would consent to participate. Some participants were seen as a dyad (mother-child), women only (child might have died or sick at the time of appointment) or child only (with caregiver). In some cases, a few participants were given an appointment but were not seen at the visit (no show).

Figure S2a: Participants retention in PRECISE-DYAD study in Kenya
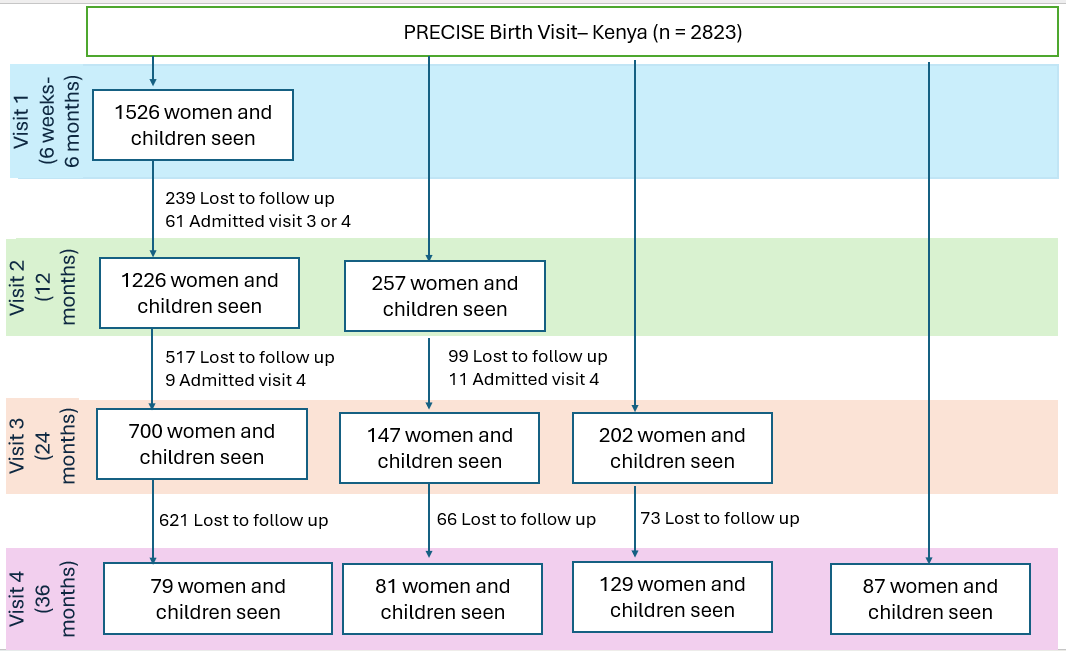

Figure S2b: Participants retention in PRECISE-DYAD study in The Gambia

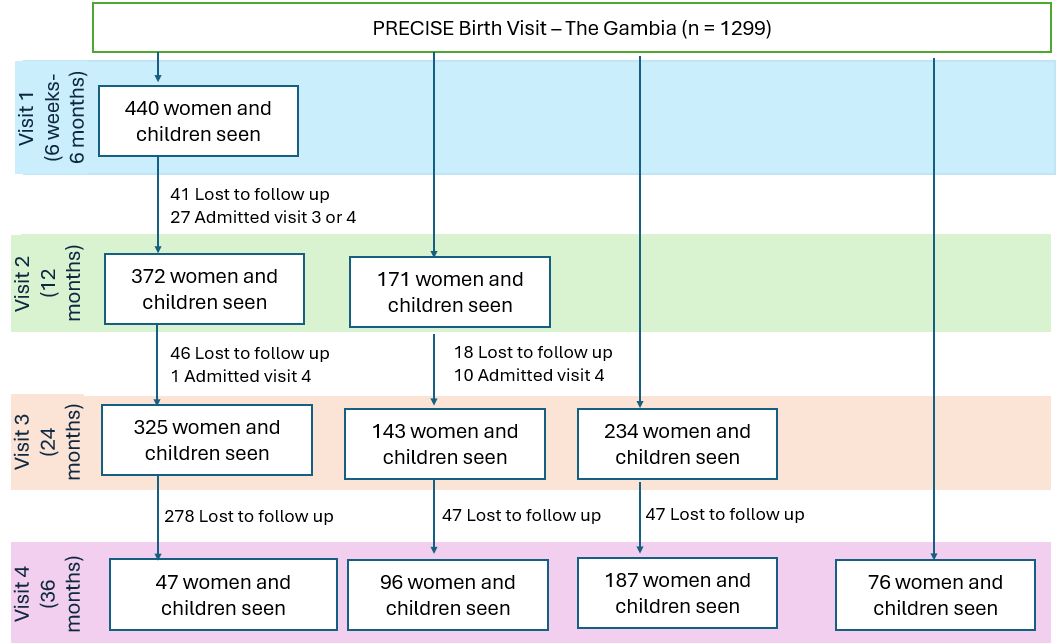

Flow chart illustrating participant entry points across the four study visits and their retention over time. Most participants enrolled at the first visit, with smaller groups joining at subsequent visits. A subset of those entering early completed all follow-up visits, while others were enrolled later in the study or attended only a single visit
